# “Sadly I think we are sort of still quite white, middle-class really” – Inequities in access to bereavement support: Findings from a mixed methods study

**DOI:** 10.1101/2022.05.12.22274992

**Authors:** L. E. Selman, E. Sutton, R. Medeiros Mirra, T. Stone, E. Gilbert, Y. Roulston, K. Murray, M. Longo, K. Seddon, A. Penny, C.R. Mayland, D. Wakefield, A. Byrne, E. Harrop

**Affiliations:** University of Bristol, Palliative and End of Life Care Research Group, Population Health Sciences, Bristol Medical School, Canynge Hall, 39 Whatley Road, Bristol BS8 2PS, U.K.; University of Bristol, Palliative and End of Life Care Research Group, Bristol, UK; Cardiff School of Dentistry, Cardiff University, Cardiff. UK; Ubele Initiative, London, UK; Cardiff University, Marie Curie Research Centre, Cardiff, UK; Wales Cancer Research Centre, Cardiff, UK; National Bereavement Alliance, London, UK; University of Sheffield, Department of Oncology and Metabolism, Sheffield, UK; North Tees and Hartlepool NHS Foundation Trust, Stockton-on-Tees, UK

## Abstract

**Background:** Voluntary and community sector bereavement services play are central to bereavement support in the UK.

**Aim:** To determine service providers’ perspectives on access to their support before and during the COVID-19 pandemic.

**Design:** Mixed methods study using an explanatory sequential design: (1) Cross-sectional online survey of UK bereavement services; (2) Qualitative interviews with staff and volunteers at case study services.

**Settings/participants:** 147 services participated in the survey; 24 interviews were conducted across 14 services.

**Results:** 67.3% of services reported there were groups with unmet needs not accessing their services before the pandemic; most frequently people from minoritised ethnic communities (49%), sexual minority groups (26.5%), deprived areas (24.5%) and men (23.8%). Compared with before the pandemic, 3.4% of services were seeing more people from minoritised ethnic groups, while 6.1% were seeing fewer. 25.2% of services did not collect ethnicity data. Qualitative findings demonstrated the disproportionate impact of the pandemic on minoritised ethnic communities, including disruption to care/mourning practices, and the need for culturally appropriate support. During the pandemic outreach activities were sometimes deprioritised; however, increased collaboration was also reported. Online provision improved access but excluded some. Positive interventions to increase equity included collecting client demographic data; improving outreach, language accessibility and staff representation; supporting other professionals to provide bereavement support; local collaboration and coproduction.

**Conclusions:** Service providers report inequities in access to bereavement support. Attention needs to be paid to identifying, assessing and meeting unmet needs for appropriate bereavement support. Identified positive interventions can inform service provision and research.

## What is already known about the topic?

- There are known inequities in who receives formal bereavement support, with, among others, people from minoritised ethnic communities, sexual minority groups and people with lower socio-economic status known to experience barriers to access.
- The COVID-19 pandemic had a disproportionate impact in the UK, with higher mortality and bereavement rates in minoritised ethnic minority communities and groups with lower socio-economic status.

## What this paper adds

- 67.3% of voluntary and community sector bereavement services in the UK reported that there were population groups with unmet support needs which experienced barriers to accessing their service before the pandemic, with minoritised ethnic groups most frequently recognised in this regard.
- Despite the disproportionate and multi-dimensional impact of the pandemic on minoritised ethnic communities, for the majority of bereavement services in the UK, the proportion of clients from minority communities did not increase and in some cases decreased during the pandemic.
- Positive interventions to increase equity included monitoring client characteristics to identify gaps; improving outreach, language accessibility and staff representation; supporting other professionals in the community to provide bereavement support; local collaboration and coproduction of services to ensure appropriateness and inclusivity.

## Implications for practice, theory or policy

- More needs to be done to tackle inequity in access to bereavement support – and many service providers both recognise this and are ready to implement changes to widen access to their support.
- Prioritising equity means identifying, assessing and meeting unmet needs in bereaved communities, adapting services and outreach to ensure inclusivity, and working in partnership with community-based organisations.
- Study findings can help inform efforts to widen access and reduce inequities.

## Background

A direct cause of approximately 6 million deaths to date, COVID-19 has brought bereavement support centre-stage as a core element of health and social care provision^1^. Public health approaches to bereavement recommend a tiered approach based on level of need^2, 3^. Tier 1 includes universal access to information on grief and available support, recognising that many bereaved people cope without formal intervention, drawing on their existing social networks. Tier 2 includes individual and group-based support for those with moderate needs, who have been shown to benefit from increased social support and opportunities for reflection, emotional expression and restorative activities. Tier 3 specialist mental health and psychological support is effective for those with high-level needs and at risk of prolonged grief disorder and should be targeted at those identified as meeting these criteria^2, 4, 5^.

In the UK, voluntary and community sector bereavement services play a crucial role in providing tier 2 and 3 bereavement support. Bereavement sector policies^6-10^ mandate equitability and fair access, yet there is evidence that certain population groups are less likely to proactively seek out and access professional care and support – even when needed and wanted – and are more likely to feel uncomfortable asking for help^11, 12^. A systematic review identified barriers to accessing bereavement support among LGBTQ+ communities as well as additional stressors, including discrimination, homophobia, disenfranchisement, historical illegality and higher rates of social isolation^13^. Another systematic review highlighted access barriers among minoritised ethnic communities: experiences of institutional racism (including in healthcare), a lack of awareness of bereavement support (often due to poor information provision by professionals^14^ and a lack of ‘in-reach’ by services), the type and/or format of support being culturally or individually inappropriate, and stigma regarding mental health within some minoritised communities^15^. Both reviews also found a lack of evidence to inform how bereavement services can best support diverse communities and ensure equitable access.

These findings are even more concerning given the disproportionate impact of the COVID-19 pandemic on people with lower socio-economic status^16^ and minoritised ethnic groups^17, 18^, reflecting underlying social, structural and economic inequalities^19^, and of an inequitable response by palliative care providers^20^. We aimed to contribute to the evidence base for equitable bereavement support by describing access to voluntary and community sector bereavement support in the UK, as reported by these organisations, and exploring bereavement service providers’ views and experiences of providing support during the COVID-19 pandemic.

## Methods

### Design

A pragmatic, explanatory sequential mixed methods study^21^ comprising:

- An online cross-sectional open survey of voluntary and community sector bereavement services in the UK, disseminated via national organisations, networks and social media (March-May 2021).
- Qualitative semi-structured telephone interviews with staff/volunteers at case study bereavement services (June-December 2021) which aimed to expand on the survey findings.

Here we present findings related to the equitability of bereavement support, using the Checklist for Reporting Results of Internet E-Surveys^22^ in reporting. This work is part of a larger research study which also examined experiences of bereavement during the pandemic in the UK^14, 23-25^

### Sampling and recruitment

Survey: A link to a JISC^26^ survey was disseminated to a convenience sample of voluntary and community sector bereavement services, via emails from the research group and national bereavement organisations and associations, national stakeholder webinars, and social media, and posted to the study website (covidbereavement.com). We asked one representative from each organisation to participate, consulting with colleagues as needed.

#### Qualitative case studies

We purposively sampled case study organisations from the 147 organisations who completed the online survey, to ensure a diverse range of services were included, considering: organisation size; geographical area; type of support provided; support for specific groups (e.g. minoritised ethnic communities, children and young people); reported challenges and innovations during the pandemic. Potential participants were sent an invitation, information sheet and consent form. If judged necessary based on the data collected and the nature of the organisation, additional staff/volunteers were recruited via snowball sampling. We also recruited participants providing support to people bereaved during the pandemic via social media communities, as these were an important source of support which was not captured in the survey. All participants gave written consent.

### Data collection

#### Survey

The survey (Supplementary file 1) comprised non-randomised open and closed questions exploring the impact of the pandemic on bereavement services and their response, including closed and open questions on access, with additional information specifically requested about clients from minoritised ethnic communities. Survey items were based on the literature and initial scoping of the pandemic’s impact^27, 28^, with input (including testing) from an expert advisory group of researchers, clinicians, bereavement support practitioners and people with experience of bereavement.

#### Qualitative care studies

Telephone interviews were conducted using a semi-structured topic guide (Supplementary file 2; adapted for online services), developed as above. Interviews were conducted by ES (n=21), EG (n=2) and LS (n=1), experienced qualitative researchers. Fieldnotes were taken to inform sampling, data collection and analysis.

### Analysis

#### Survey

All data are categorical. Graphical summaries, including pie charts, bar charts and stacked bar charts, were used to describe all variables. Logistic regressions were performed to investigate which factors (area served, type of organisation, client group/age, whether restricted by cause of death or age of deceased) might be associated with reporting they were not reaching specific community groups with unmet needs; the proportion of clients from minoritised ethnic groups; and whether the organisation collected ethnicity data. All analyses were performed by RM using R (version 4.1.1, R Core Team, 2021), implemented in R-Studio (www.r-studio.com). Free-text data were analysed using thematic analysis^29, 30^ in NVivo12^31^ by TS, discussed with LS and ES and refined.

#### Case studies

Interviews were transcribed verbatim and checked for accuracy prior to thematic analysis^29, 30^ in NVivo12^31^. Analysis used a combination of deductive and inductive coding strategies and was conduced concurrently with data collection, allowing insights from earlier interviews to inform those conducted subsequently. ES, LS and TS read and independently coded a sub-set of interview transcripts and developed a coding framework which ES applied to the dataset. ES and LS met regularly to discuss the development and revision of key themes and sub-themes^32^.

Quantitative and qualitative findings were triangulated and integrated into a narrative, with the latter used to explain and add richness to quantitative findings^21^. All quotations are anonymised.

### Ethical approval

Ethical approval for the study was granted by the University of Bristol, Faculty of Health Sciences Ethics Committee (Ref: 114304 20/12/2020).

## Results

### Participants

#### Survey

147 bereavement services from across UK regions participated (Figure 1). Two participants completed the survey twice; their first and second responses were merged. Two services provided two responses; the second response from each was excluded. 44.5% were hospice or palliative care services (including services part-funded by the NHS); 15.1% national bereavement charities or non-governmental organisations (NGOs); 11.6% local bereavement charities/NGOs; 8.9% branch of a national bereavement charity/NGO; 4.1% branch of other national charity/NGOs; 6.8% other local charities/NGOs; 8.9% other (e.g. council-commissioned service, local collaborative partnership, community-led initiative or community interest company). 68% provided support following all causes of death whereas 32% were focused on specific causes of death such as terminal illness.

**Figure 1:**
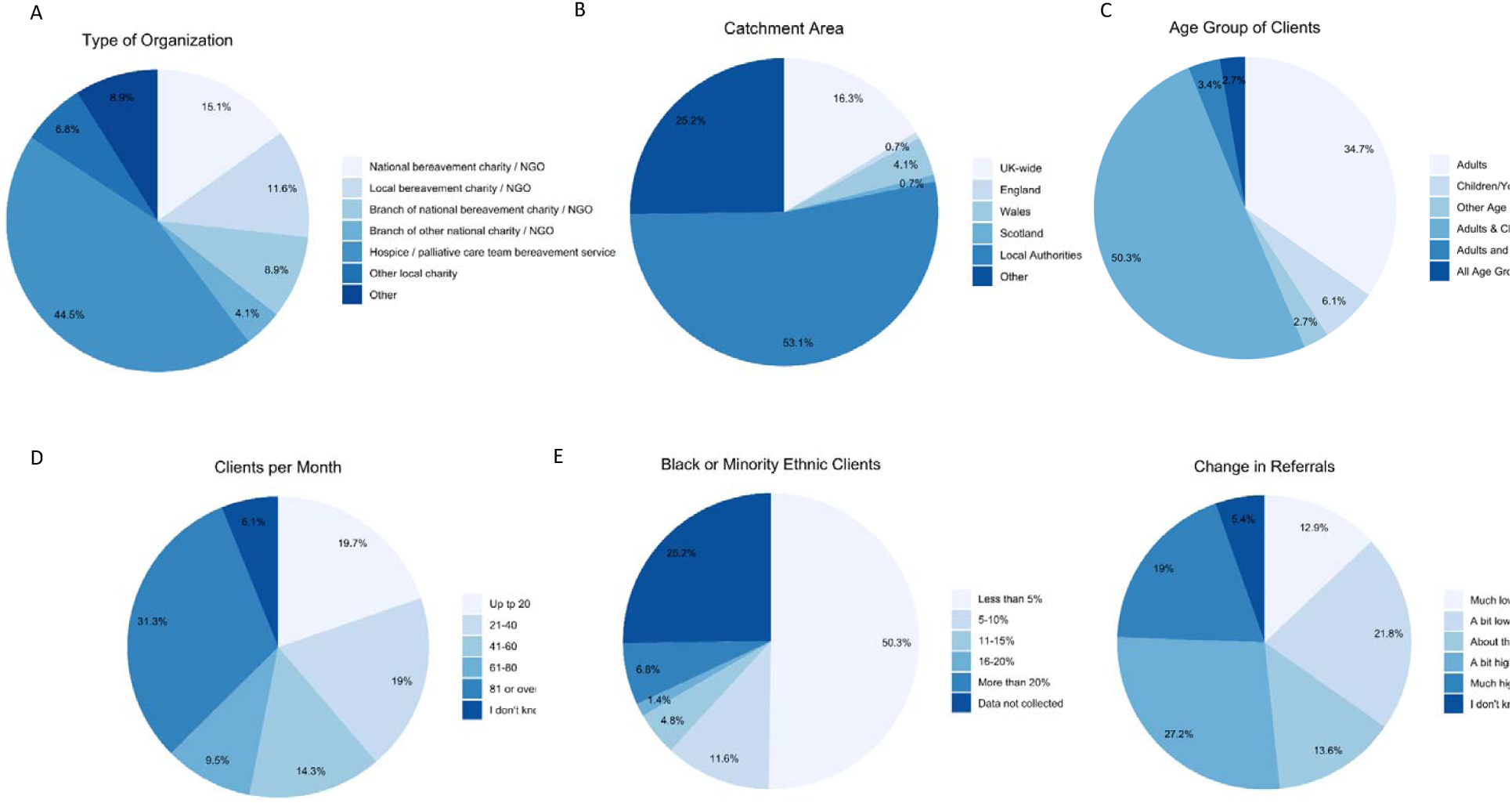
Characteristics of service and change in referrals (n=147, except for Type of Organisation n=146)

#### Case studies

Twenty-four interviews with staff and volunteers from 14 organisations were conducted (Table 1). Two services provided targeted support for specific minoritised ethnic communities (Muslim and African Caribbean). Fourteen survey respondents were sampled and approached via email; 3 did not respond. A further 12 participants were suggested via snowball sampling; 2 did not respond. Two potential participants coordinating social media communities were contacted via email and both participated; an additional volunteer was recruited via snowball sampling. Interviews lasted 25-77 minutes (mean 46 mins).

**Table 1:** Details of case study organisations and interview participants

| **Organisation ID** | **Size of Org** | **Specialist support provided** | **Geographical area** | **Interviewee role** |
| --- | --- | --- | --- | --- |
| **Org A** | Regional | Hospice, family support | South East | **A1:** Manager<br>**A2:** Director |
| **Org B** | Small | Minoritised group (Muslim community) | UK wide | **B1:** Director<br>**B2:** Volunteer |
| **Org C** | Branch of National Organisation | General bereavement support | West Midlands | **C1:** Regional Manager<br>**C2:** Volunteer |
| **Org D** | Small | Minoritised group (African Caribbean community) | UK Wide | **D1:** Director |
| **Org E** | Regional | Hospice, Regional Bereavement Network | Scotland | **E1:** Team Lead<br>**E2:** Volunteer |
| **Org F** | Small, Regional | Children and young people, Bereavement Network | Scotland | **F1:** Coordinator |
| **Org J** | Regional | Counselling, individual and group therapy | South West | **J1:** Clinical Lead<br>**J2:** Senior Therapist |
| **Org K** | Regional | Hospice, family support | North East | **K1:** Senior Practitioner<br>**K2:** Volunteer |
| **Org P** | Branch of National Organisation | Palliative care, counselling | South East | **P2:** Specialist Counsellor |
| **Org N** | Small, Regional | Hospice, family support | Wales | **N1:** Team Lead<br>**N2:** Social Worker |
| **Org M** | Branch of National Organisation | General Bereavement Support | Northern Ireland | **M2:** Regional Manager<br>**M3:** Volunteer |
| **Org Q** | National Organisation | Telephone Support | UK Wide | **Q1:** Coordinator<br>**Q2:** Volunteer |
| **Org H** | Online Organisation | COVID-specific, mutual support | UK Wide | **H1:** Founder |
| **Org L** | Online Organisation | COVID-specific, mutual support | UK Wide, Wales | **L1:** Founder<br>**L2:** Regional Administrator |

### Groups with unmet needs

67.3% of services reported that there were groups with unmet needs which were not accessing their services before the pandemic. The most frequently recognised of these was people from minoritised ethnic communities (n=72, 49% of total), followed by sexual minority groups (26.5%), socio-economically deprived communities (24.5%), men (23.8%) and ‘other’ (15%) (including digitally excluded, homeless people, people with learning disabilities, travelling community, non-English speakers, rural communities, physically disabled or with mobility problems). Most organisations that reported being unable to reach certain community groups were not reaching two or more specific groups (71% of those reporting difficulties reaching specific community groups and 48% of the total) (Figure 2). None of the variables used were significant in predicting which organisations were more likely to report being unable to reach specific groups (Supplementary file 3).

**Figure 2:**
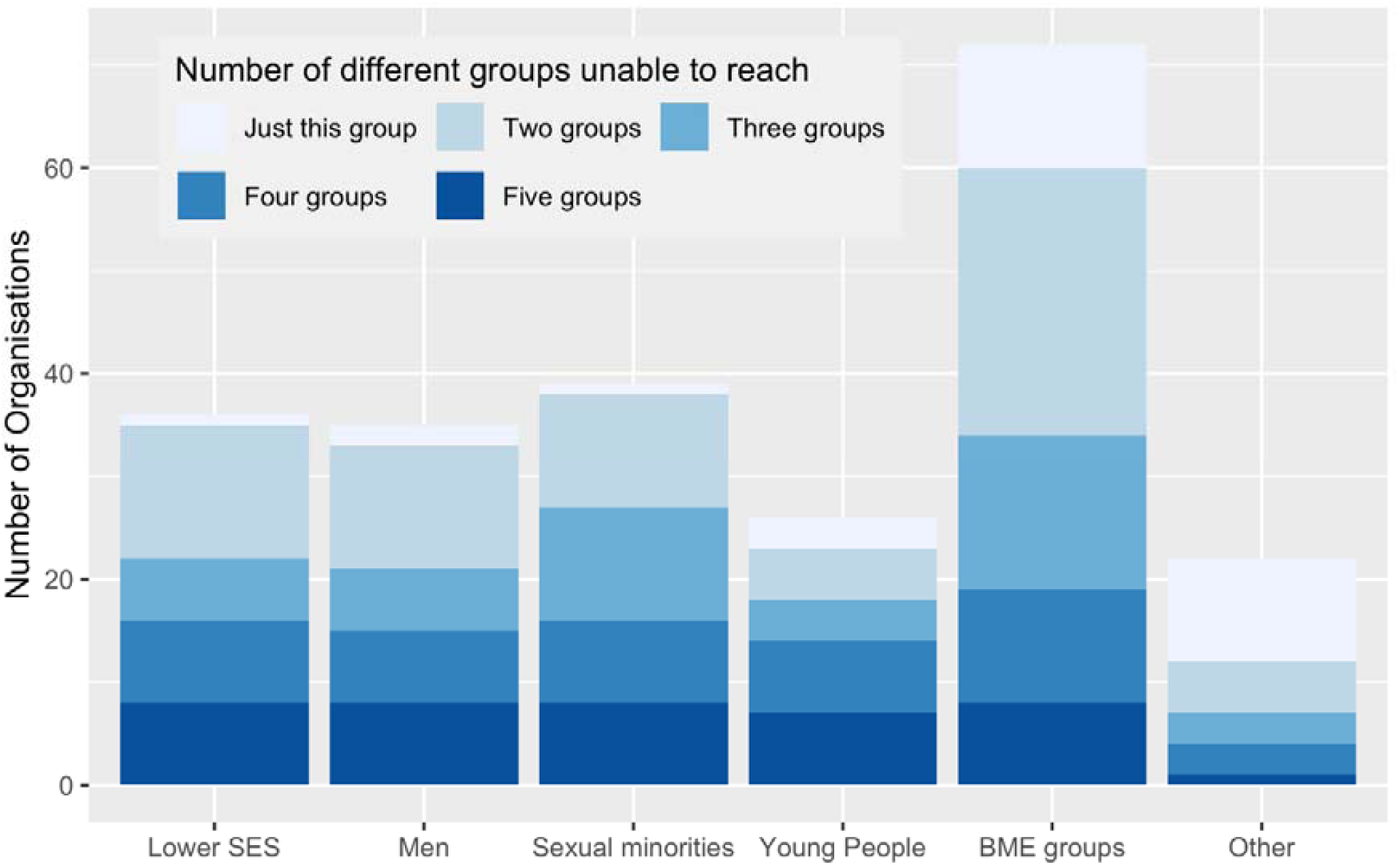
Specific community groups with unmet bereavement support needs not being reached/experiencing access barriers (N=98) SES = socio-economic status; BME = Black and minoritised ethnic Note: Answers to question 15a: If you selected ‘Yes’ [to question 15: Before COVID-19, do you think there were specific community groups with unmet bereavement support needs that you were not reaching, or who experienced barriers to accessing your service?)], which groups? Please tick all that apply.

The recognition that bereavement support was not equitable was reflected in qualitative data:

> *“one of the big issues that we face as an organisation is actually being able to reach Black and ethnic minority populations, lower socio-economic groups. We really struggle to reach them. You know sadly, well it’s not sad for those people that come to us, but sadly I think we are sort of still quite white, middle-class really.” (J1, Regional organisation)*
>
> *‘I don’t have figures to back this statement up, but from observations, it appears that the client group we are reaching tend to be from a middle class background, they are more educated and have a greater awareness of the support available in the community. We have less referrals from those from a lower socio-economic background.’ (Survey ID122, Branch of national charity/NGO (not bereavement-specific))*

### Access by minoritised ethnic communities

50.3% of organisations reported that, in the year before pandemic, <5% of their clients were from minoritised ethnic groups, while 6.8% of services (predominantly London-based) reported that >20% of their clients were from minoritised ethnic groups; 25.2% reported not collecting ethnicity data (Figure 1). 45% of those organisations with less than 5% of their clients from minoritised ethnic groups did not report that those communities had unmet needs for their support.

There was a trend towards an increasing number of clients (i.e. larger organisations) being associated with an increase in the odds of reporting ≥ 5% clients from minoritised ethnic groups, but the differences in odds were only significant between the largest organisations and the smallest ones: organisations with ≥80 clients per month were 3.8 and 7 times more likely to have ≥5% clients from minoritised ethnic groups compared to organisations with 21-40 clients per month and ≤20 clients per month, respectively (Figure 3 and Supplementary file 3).

**Figure 3:**
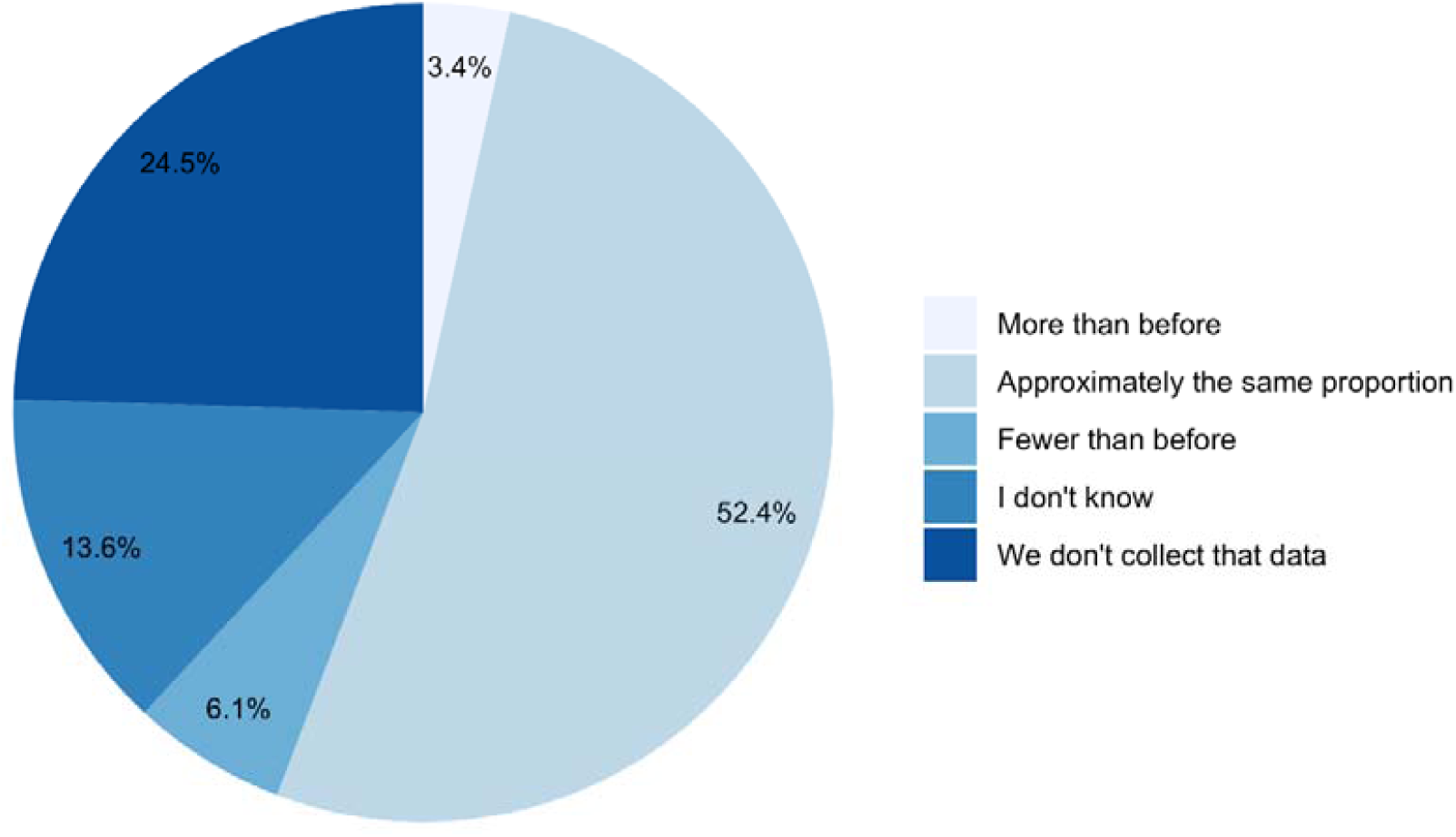
Change in proportion of clients from minoritised ethnic communities since before the COVID-19 pandemic (n=147)

**Figure 4.**
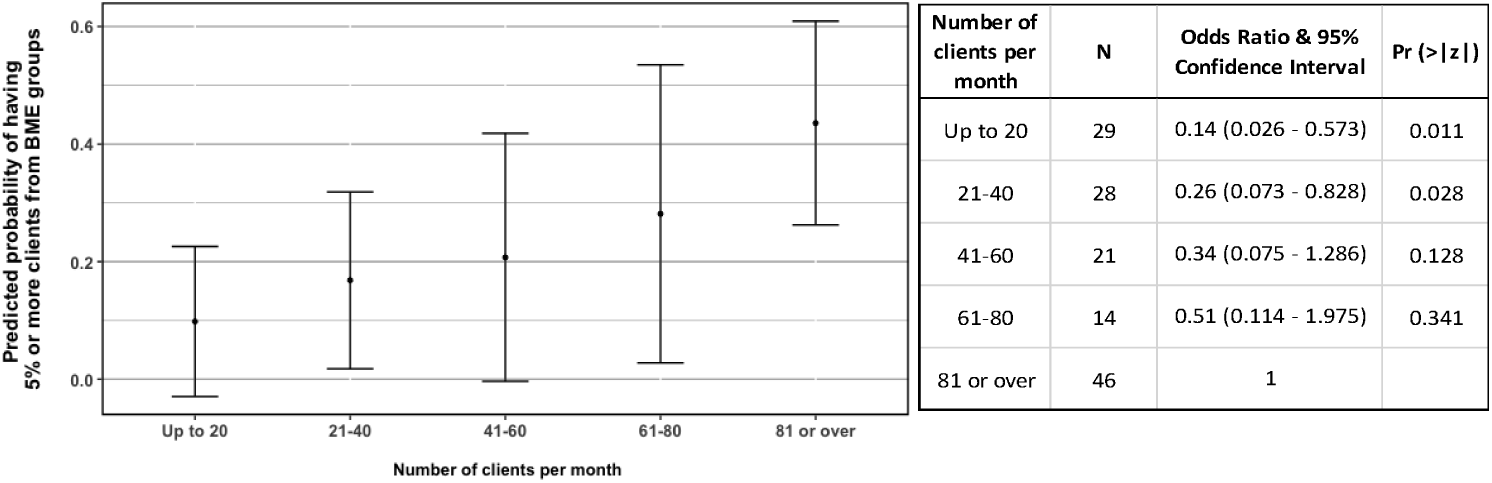
Left - Probabilities of an organisation having 5% or more clients from Black and minoritised ethnic (BME) groups in relation to number of clients per month, predicted from the Logistic Regression parameters (Supplementary file 3). The final model where predictions were calculated from also included the variable “focus on children and young people”, which was set to “yes”, as the best represented category. Right – Table of Odds Ratios and 95% Confidence Intervals for Number of clients per month (reference category: 81 or over)

There are apparent regional differences in the proportion of clients from minoritised ethnic groups across organisations (Table 2), but catchment area (UK-wide, nation-specific, county or locally-specific, or other) was not a significant predictor in the analysis. Region of the UK could not be used in the analysis due to very small sample sizes across most regions for services with ≥5% ethnically minoritised clients (Table 2). There was a possible relationship between organisations supporting primarily children or young clients being more likely to collect information on the ethnicity of their clients (OR = 1.84, 95% CI = 0.938 – 3.652), but there is a relatively high probability that this result could be due to chance (p=0.077; Supplementary file 3), hence this finding should be considered with caution.

**Table 2:** Contingency table heat-map relating region of the UK served by the organisation with responses to the question “In the year before COVID-19, what proportion of your clients were from Black or minority ethnic communities? Please select your closest estimate.” Light-to-dark shading represents increasing numbers of organisations (n=147)

| Region of the UK served | Proportion of clients from Black and minoritised ethnic communities |  |  |  |  | We don't collect this data | Total which collected this data | Total |
| --- | --- | --- | --- | --- | --- | --- | --- | --- |
|  | Less than 5% | 5-10% | 11-15% | 16-20% | More than 20% |  |  |  |
| UK (all) | 5 | 3 | 2 |  | 2 | 12 | 12 | 24 |
| England (only) |  |  |  |  | 1 |  | 1 | 1 |
| Wales (only) | 8 | 3 |  |  |  | 3 | 11 | 14 |
| Scotland (only) | 6 | 1 |  |  |  |  | 7 | 7 |
| NI (only) | 2 |  |  |  |  |  | 2 | 2 |
| London (greater London) |  | 1 |  | 1 | 5 |  | 7 | 7 |
| South East | 10 | 3 | 2 |  |  | 11 | 15 | 26 |
| East of England | 6 | 1 | 1 |  |  | 2 | 8 | 10 |
| East Midlands | 4 | 1 |  |  |  | 1 | 5 | 6 |
| North East | 2 | 1 |  |  |  | 1 | 3 | 4 |
| North West | 10 |  |  |  |  | 3 | 10 | 13 |
| West Midlands | 4 |  | 1 |  | 2 | 2 | 7 | 9 |
| South West | 8 | 1 | 1 | 1 |  |  | 11 | 11 |
| Yorkshire & the Humber | 5 | 2 |  |  |  | 2 | 7 | 9 |
| East Midlands AND South East | 1 |  |  |  |  |  | 1 | 1 |
| North West AND Wales | 1 |  |  |  |  |  | 1 | 1 |
| Wales AND West Midlands | 1 |  |  |  |  |  | 1 | 1 |
| Yorkshire & Humber AND East Midlands | 1 |  |  |  |  |  | 1 | 1 |

There was variation in how referrals overall had changed during the pandemic, with 35% of services reporting lower numbers compared with before the pandemic and 46% reporting higher numbers (Figure 1). Compared with before the pandemic, 3.4% of services were seeing more people from BME groups, 6.1% were seeing fewer, and 38% either didn’t know or didn’t collect this data (Figure 3).

The variation in access by people from minoritised ethnic communities was reflected in qualitative data, e.g.

> *Massive increase in access by our South Asian and African Caribbean communities - Early access - more people are coming to us within the first 6 months of their bereavement - Covid-19 is the second highest cause of death - 60% increase in access - compared to same 12 months last year.’ (Survey ID94, Branch of national bereavement charity/NGO)*
>
> *We’ve had so many less referrals from ethnic minorities than normal, yeah, and if we’ve had, if someone’s identified themselves from an ethnic minority background, they’ve usually got really good English, whereas before we have had to use our interpreting service or some other way of talking to people. (A1, Hospice)*

### Disproportionate impact and cultural appropriateness

Participants from services supporting minoritised ethnic communities described the multidimensional and acute impact of the COVID-19 pandemic on their clients, in terms of the number and nature of deaths but also the disruption to caring practices:

> *‘We think that the increase in minority groups contacting us is directly related to the numbers of the BAME [Black, Asian and minoritised ethnic] community dying from Covid-19. Very sadly, some of the callers to our service have had almost whole families wiped out from Covid-19*.*’ (Survey ID44, National bereavement charity/NGO)*
>
> *In terms of the way some families work and the dynamics of them; there’s real emphasis on caring for your own elders and all of that, again, has just been eliminated by the fact that these deaths are often – in fact, the vast majority – in hospital wards and because of the restrictions it’s not possible to go and see your loved one until you get that call to say they’re dying… so it’s sudden, it’s unexpected, there isn’t that closure. Afterwards, too, the support that you tend to get after from the wider community visiting your home and offering their consolation and things like that, that’s all absent as well. (B1, Small organisation for minoritised ethnic groups)*

This disruption also extended to mourning traditions, as infection control restrictions had a disproportionate impact on communities with strong and important rituals involving cleansing or viewing the deceased’s body or coming together in large groups to support the bereaved:

> *From the Muslim perspective… there are certain rites when it comes to burial and death*… *It might sound very odd to somebody who doesn’t share those beliefs, but one of the things that is really important is to be part of that burial process, to wash and shroud the deceased loved ones of yours. It’s almost as though there’s a pride, almost, for that individual to give them away to what we believe is the next world, if you like – we believe in the hereafter, life after death – and the closure that that would’ve resulted in has gone away into the ether*.. *(B1, Small organisation for minoritised ethnic groups)*
>
> *‘BAME [Black, Asian and minoritised ethnic] communities have expressed interruption of spiritual and cultural rituals of marking the passing of a loved one in all diverse communities*.*’ (Survey ID32, National bereavement charity/NGO)*

These particular challenges in bereavement highlighted the importance of cultural appropriateness in bereavement support and the role of religious support and guidance:

> *We also offer support from a religious perspective… ‘Where is my child now…?…If I were to visit their grave, would they be able to hear me?’, those sorts of questions are really quite important to grief processes [and]… can only really be answered by somebody who has an insight into those things. We don’t always have all the answers… but we do have access to imams and resources… over time we’ve accumulated a database of commonly-asked questions. (B1, Small organisation for minoritised ethnic groups)*

Understanding and accommodating cultural traditions was essential; for example, a participant from a service focused on supporting African Caribbean communities contrasted a family’s poor experiences at a national charity with their own understanding and accommodation of African Caribbean traditions such as 9 Nights:

> *I have heard stories, not to pick on [National Organisation], but I have heard stories, family that came to us said they didn’t have a good experience*… *We can go into things you know, we have our references, how we do things… we do our wakes, what we call 9 Nights and all the traditional things that we can also include in our support*… *We understand that expression. (D1, Small organisation for minoritised ethnic groups)*

Finally, specific cultural stressors which need to be understood by bereavement support providers were described; for example, how close community support can sometimes present its own challenges:

> *In the Muslim community we tend to have very much a community spirit and we do tend to help one another but sometimes that can be quite suffocating … it’s really hard to confide in people without making them upset or making them feel more hurt than they already are… so you tend to kind of bottle things up… [but] you’re still carrying the grief aren’t you. (B2, Small organisation for minoritised ethnic groups)*

### Effects of the pandemic on access

Participants reported a huge move to online services, which had both positive and negative impacts on accessibility. On the one hand, it reduced waiting times and improved reach and accessibility for groups who might have been excluded by face-to-face services, for example, carers, those in rural communities and Muslim women:

> *Muslim women tend to go through something called the Iddah period after they lose their husband where, for four months and 10 days, it’s a period of reflection where they tend to stay at home, and for them to access this support is now a possibility. (B1, Small organisation for minoritised ethnic groups)*

It also benefitted groups who preferred online provision – men and young people were mentioned in this regard, e.g.

> *Yes - the remote nature and greater use of social media has helped to engage a younger population*.*’ (Survey ID119, National bereavement charity/NGO)*

With the use of online services, access to support could also be faster and less restricted – for example, by where volunteers were based. However, some people were excluded, particularly people already experiencing disadvantage, for example, due to illiteracy or a lack of access to technology:

> *Having these session via Zoom will exclude some people but when I’m asking the triage team on our helpline what percentage of people does it exclude, they would say about 10-15%, so the majority of people [can access it], even our older community*… *It tends to be people with maybe very serious mental health conditions or on the peripheries of society or who are digitally excluded obviously can’t attend. (C1, Branch of Large National Organisation)*
>
> *We’ve got an illiteracy rate of something like 20 percent in [area] so our client group are already disadvantaged by poor reading and writing skills irrespective of their technological skills. (N1, Hospice)*

Other groups reported to find online support difficult or impossible included young children, those who required text speak or translation services, and parents and carers with childcare responsibilities.

Participants described other ways the pandemic had affected services’ ability to meet the needs of diverse groups. First, in the context of uncertainty and lack of resources, outreach activities had sometimes been deprioritized or impossible:

> *However, myself and other colleagues used to go out and promote services face to face to BAME [Black, Asian or minoritised ethnic] groups and obviously this hasn’t happened. (Survey ID2, Hospice)*
>
> *We have always had our hard-to-reach communities and I think they have potentially become harder to reach because they closed off during COVID, so we would want to start to engage that work again… We are just doing a piece of work around palliative care and learning disabilities and we would like to look at homelessness and our travelling community… there is lots of scope to re-invigorate work that was already in place that got pulled*. (A2, Hospice)

Second, the lack of capacity in mental health services had knock-on effects, with a participant commenting on the lack of follow up from specialist “crisis” teams:

> *There have been struggles or perhaps capacity issues with people when for instance we’ve contacted Crisis teams and they haven’t necessarily followed up on individuals. So I get the sense… that it’s been quite stretched or difficult in some areas. (Q1, Large National Organisation)*

### Positive interventions

Service providers described positive interventions they had implemented or wanted to implement to try to reduce inequity of access. A fundamental step was capturing clients’ demographic data to understand who was and wasn’t accessing the service:

> *We do not capture data on sexuality, race etc. for all of our clients at the beginning of accessing our support, this is only captured at the end and is optional (so we don’t have data that represents the majority of people accessing our support). We are looking at better ways to be capturing this data so it gives a more accurate representation*.*’* (Survey ID73, Branch of a national charity/NGO)

Through collaboration with community groups and other organisations as well as GPs, services aimed to improve referral pathways, signposting, outreach and advertising to specific communities:

> *‘We have partnered with [Black women’s group] and this has developed engagement from BAME [Black, Asian or minoritised ethnic] communities*.*’ (Survey ID139, National bereavement charity/NGO)*
>
> *‘It has been positive to work with other local charities and organisations. Connections not only local, but Twitter/Facebook have enabled wider contact, sharing of knowledge, reduction in barriers’. (Survey ID80, Community-led peer support)*

Attending to language and representation was seen a key way of improving access:

> *“On our triage team, one of my colleagues, Imrat, she speaks Punjabi and Urdu, we’ve recruited a lot more volunteers as well that speak various languages, so it’s making us more accessible, more diverse, which is amazing. Anita, who also answers the triage line, she’s from the West Indies, she’s got a very distinct accent, and do you know what, it makes callers feel very, very comfortable*… *just to have a diverse range of people answering the triage line is incredible”* (C2, Branch of Large National Organisation; pseudonyms used)

Some organisations were engaged in projects to try to reach marginalised groups, had introduced new services to target perceived gaps in their provision, and/or reported supporting other professionals (e.g. community workers) to provide bereavement support themselves. Other activities included engaging an external organisation to look at equality and diversity across an organisation, and designing and implementing an organisational strategy focussed on inclusivity, diversity and outreach; for example:

> *Equality and diversity is a big theme in [National Organisation] at the minute, and we’ve actually engaged an organisation to have a look at all our stuff and see how, from an objective point of view, how it all looked in terms of equality and diversity I have linked in with groups… that support LGBTQ communities… about… what makes us as an organisation approachable to your clients… they need to know who they’re going to for support isn’t going to be shocked or uncomfortable with a same sex relationship. So, for us it’s about, how do we promote [National Organisation] as an organisation that is friendly and supportive for all, race, sex, religion, whatever? (M2, Branch of national organisation)*

## Discussion

### Main findings

Two thirds of UK voluntary and community sector bereavement services recognised that there were population groups which could benefit from their services, but do not access them. People from minoritised ethnic groups were most frequently recognised in this regard, followed by sexual minority groups, people with lower socio-economic status and men. During the pandemic, on average, proportions of ethnically minoritised clients did not increase despite the disproportionate multidimensional impacts of the pandemic on these communities – in terms of mortality rate and care and mourning practices, but also compounding factors such as social and economic inequity, racism and discrimination.

### What this study adds

One approach to widening access was to expand advertising and focus on ‘in-reach’, for example via other community organisations or groups. Proactive advertising via local community networks and organisations was an important feature of the immediate disaster response to the 911 attacks in New York, which achieved high uptake of counselling support among minoritised ethnic groups^33, 34^. Similarly, interventions which focus on language and visibility, which we also identified, can help encourage engagement with formal bereavement support^5, 33^.

However, previous studies have highlighted cultural inappropriateness in bereavement support^35, 36^. If services are, albeit unintentionally, inappropriate, insensitive or biased, then raising awareness of services among disadvantaged communities, providing linguistic access or ensuring a diverse workforce will not, on its own, create equity. Appointment of bilingual health-care workers can help bring about better family support both before and after the death^37^, yet it is their awareness of cultural proprieties around death and mourning which is likely crucial to their success.

More extensive interventions to widen access to bereavement support which we identified therefore examined how organisational structures and approaches to care could exclude diverse groups from engaging with or benefitting from bereavement services and adapted services accordingly. These kinds of interventions are often based on consultation or co-production with disadvantaged groups, prioritise cultural competence and service adaptation, and may involve collaboration with other organisations with specific cultural, faith, legal or financial expertise to help meet group-specific support needs. Systematic reviews have established the importance of cultural knowledge and sensitivity in bereavement support following mass-bereavement events^33^ and in palliative care^5^. A recent survey of mental health services for minoritised ethnic communities in the UK similarly identified a need for mainstream therapists and service providers to have quality-assured cultural competency training^12^. The survey also identified increased demand for bereavement support provided by organisations led by people from minoritised ethnic commmunities during the pandemic^12^, highlighting the importance of supporting community organisations representative of and trusted by the populations which they serve.

### Limitations

We did not collect detailed data on the range of groups not accessing services or how these might have changed during the pandemic. Our decision to focus on minoritised ethnic groups was informed by evidence of the impact of the pandemic at the time of the survey and pragmatic concerns regarding participant burden. Research into access to bereavement services by other disadvantaged groups is crucial given what is now known regarding the pandemic’s impact^38^. Convenience sampling might have resulted in less burdened or more engaged services completing the survey. It is not known precisely how many voluntary and community sector bereavement services there are in the UK; a 2020 analysis of services registered on a national directory identified 822 entries^28^, however this is likely to include services outside the sector and services no longer operating. For practical reasons the survey considered marginalised groups as separate entities rather than intersecting in complex ways; study interviews with bereaved people will explore access and intersectionality in more depth. Given diversity in types of catchment area, it was not possible to conduct a detailed analysis of proportions of ethnically minoritised clients in services serving distinct regions of the UK and compare these with local population characteristics.

### Implications and recommendations

#### On the basis of study findings, we recommend that

- Collection of client data on all key protected characteristics^6, 39^ and outcomes of support becomes routine in all bereavement services, to enable a better understanding of access, ‘reach’ and effectiveness, to help ensure equity and meet the needs of the whole population. A quarter of services currently do not have accurate data relating to ethnicity.
- Services assess unmet needs for formal bereavement support in the local community, recognising that not everyone will need professional support but that appropriate support should be offered and available to anyone who might benefit. Client data can then be compared with local population characteristics and needs assessment, based on catchment area and target population. Within an organisation, open discussion of this data may help create internal motivation^40^ to change practice and improve equity.
- Because a ‘one size fits all’ approach will never achieve equity, service providers must ask sometimes difficult and uncomfortable questions about the nature of their service, and build on basic interventions such as in-reach, language accessibility and diversity of representation to consider how organisational structures and assumptions could preclude beneficial support.
- Implicit bias, anti-discrimination and cultural competency training should be routine for bereavement providers – and the mental health services signposting to them.
- Local knowledge, collaboration with other community-based organisations, and co-production should be standard approaches in the design and delivery of bereavement services.
- Since online services have drawbacks as well as benefits in terms of accessibility, with the most disadvantaged often the most likely to be excluded, research is needed to further understand the acceptability and outcomes of online support in different groups.
- Financial resources and support are provided to community organisations working with minoritised groups, strengthening the bereavement support that they are able to provide to the communities they serve. Bereavement services can help advise and support other community stakeholders.
- Research is conducted to identify best practice interventions to reduce inequity in access to bereavement support.

## Supporting information

Suppl file 1 Survey

Suppl file 2 Topic guide

Suppl file 4 Cherries Reporting Checklist

## Data Availability

Full study data sets will be made available in May 2022, 3 months after study closure. Data sharing requests should be directed to Dr. Lucy Selman.

## Acknowledgements

Our thanks to all the organisation representatives who completed the survey, and to all the individuals and organisations that helped disseminate the survey. We would also like to thank the project assistants, collaborators and wider advisory group members who are not co-authors on this publication: Dr. Emma Carduff, Dr Daniella Holland-Hart, Dr. Linda Machin, Dr. Anne Finucane, Prof. Bridget Johnston, Dr. Silvia Goss, Dr Kirsten Smith, Dr. Audrey Roulston, Dr. Liz Rolls and Dr. Anna Torrens-Burton.

## Financial support

The author(s) disclosed receipt of the following financial support for the research, authorship, and/or publication of this article: This study was funded by the UKRI/ESRC (Grant No. ES/V012053/1). The project was also supported by the Marie Curie core grant funding to the Marie Curie Research Centre, Cardiff University (grant no. MCCC-FCO-11-C). E.H., A.B. and M.L. posts are supported by the Marie Curie core grant funding (grant no. MCCC-FCO-11-C). ATB is funded by Welsh Government through Health and Care Research Wales. K.V.S. is funded by the Medical Research Council (MR/V001841/1). The funder was not involved in the study design, implementation, analysis or interpretation of results and has not contributed to this manuscript.

## Conflicts of interest

None

## Ethical standards

The authors assert that all procedures contributing to this work comply with the ethical standards of the relevant national and institutional committees on human experimentation and with the Helsinki Declaration of 1975, as revised in 2008.

## Notes

### Competing Interest Statement

The authors have declared no competing interest.

## References

1. Organization. WH. WHO Coronavirus Disease (COVID-19) Dashboard, https://covid19.who.int/ (2022, accessed 11.02.2021).

2. NICE. Guidance on cancer services: Improving supportive and palliative care for adults with cancer. The manual. London2004.

3. Aoun SM, Breen LJ, O’Connor M, et al. A public health approach to bereavement support services in palliative care. Aust N Z J Public Health 2012; 36: 14-16. 2012/02/09. DOI: 10.1111/j.1753-6405.2012.00825.x.

4. Aoun SM, Breen LJ, Howting DA, et al. Who Needs Bereavement Support? A Population Based Survey of Bereavement Risk and Support Need. PLOS ONE 2015; 10: e0121101. DOI: 10.1371/journal.pone.0121101.

5. Harrop E, Morgan F, Longo M, et al. The impacts and effectiveness of support for people bereaved through advanced illness: A systematic review and thematic synthesis. Palliative Medicine 2020; 34: 871–888. DOI: 10.1177/0269216320920533.

6. Welsh Government. National framework for the delivery of bereavement care 2021. Cardiff: Welsh Government.

7. Penny A and Relf M. A guide to commissioning bereavement services in England 2017. London.

8. Cruse Bereavement Care and the Bereavement Services Association. Bereavement Care Service Standards 2014. London.

9. Hall C, Hudson P and Boughey A. Bereavement support standards for specialist palliative care services 2012. Melbourne.

10. National Alliance for Children’s Grief. NAGC Standards of Practice 2013. Texas, USA.

11. Sue Ryder. A Better Grief 2019. London, UK: Sue Ryder.

12. Murray K. National Mapping of BAME Mental Health Services 2020. London.

13. Bristowe K, Marshall S and Harding R. The bereavement experiences of lesbian, gay, bisexual and/or trans* people who have lost a partner: A systematic review, thematic synthesis and modelling of the literature. Palliative Medicine 2016; 30: 730-744. DOI 10.1177/0269216316634601.

14. Selman LE, Farnell D, Longo M, et al. Risk factors associated with poorer experiences of end-of-life care and challenges in early bereavement: Results of a national online survey of people bereaved during the COVID-19 pandemic. Palliat Med 2022 Feb 17 2022.

15. Mayland CR, Powell R, Clarke G, et al. Bereavement care for ethnic minority communities: A systematic review of access to, models of, outcomes from, and satisfaction with, service provision. PLoS One 2021; 30;16(6):e0252188. DOI: 10.1371/journal.pone.0252188.

16. Naylor-Wardle J, Rowland B and Kunadian V. Socioeconomic status and cardiovascular health in the COVID-19 pandemic. Heart 2021; 107: 358–365. DOI: 10.1136/heartjnl-2020-318425.

17. Public Health England. Disparities in the risk and outcomes of COVID-19 2020. London: Public Health England

18. Office for National Statistics. Updating ethnic contrasts in deaths involving the coronavirus (COVID-19), England: 8 December 2020 to 1 December 2021 2022.

19. Scientific Advisory Group for Emergencies (SAGE). Drivers of the higher COVID-19 incidence, morbidity and mortality among minority ethnic groups, 23 September 2020 2020. London: Uk Government.

20. Bajwah S, Koffman J, Hussain J, et al. Specialist palliative care services response to ethnic minority groups with COVID-19: equal but inequitable—an observational study. BMJ Supportive & Palliative Care 2021: bmjspcare-2021-003083. DOI: 10.1136/bmjspcare-2021-003083.

21. Creswell JW and Plano Clark VL. Designing and Conducting Mixed Methods Research. 3rd ed. London: Sage 2018.

22. Eysenbach G. Improving the quality of Web surveys: the Checklist for Reporting Results of Internet E-Surveys (CHERRIES). Journal of medical Internet research 2004; 6: e34–e34. DOI: 10.2196/jmir.6.3.e34.

23. Harrop E, Goss S, Farnell D, et al. Support needs and barriers to accessing support: Baseline results of a mixed-methods national survey of people bereaved during the COVID-19 pandemic. Palliative Medicine 2021; In Press.

24. Torrens-Burton A, Goss S, Sutton E, et al. ‘It was brutal. It still is’: A qualitative analysis of the challenges of bereavement during the COVID-19 pandemic reported in two national surveys [preprint]. medRxiv 2021. DOI: 10.1101/2021.12.06.21267354.

25. Harrop E, Goss S, Longo M, et al. Parental perspectives on the grief and support needs of children and young people bereaved during the Covid-19 pandemic: Qualitative findings from a national survey. medRXiv [Preprint] 2021. Preprint 07/12/2021. DOI: 10.1101/2021.12.06.21267238.

26. JISC. JISC Online Surveys, https://www.onlinesurveys.ac.uk/ (2021, accessed 13.9.2021 2021).

27. Pearce C, Honey JR, Lovick R, et al. ‘A silent epidemic of grief’: a survey of bereavement care provision in the UK and Ireland during the COVID-19 pandemic. BMJ open 2021; 11: e046872. 2021/03/05. DOI: 10.1136/bmjopen-2020-046872.

28. Penny A and Nibloe R. Covid-19: the response of voluntary sector bereavement services. A report from the National Alliance and Childhood Bereavement Network 2021. London.

29. Braun V and Clarke V. Using thematic analysis in psychology. Qualitative Research in Psychology 2006; 3: 77–101. DOI: 10.1191/1478088706qp063oa.

30. Braun V, Clarke V and Weate P. Using thematic analysis in sport and exercise research In: Smith B and Sparkes AC (eds) Routledge handbook of qualitative research in sport and exercise. London: Routledge, 2016, pp.pp. 191–205.

31. QSR International Pty Ltd. NVivo (Version 12). 2018.

32. Braun V and Clarke V. Reflecting on reflexive thematic analysis. Qualitative Research in Sport, Exercise and Health 2019; 11: 589–597. DOI: 10.1080/2159676X.2019.1628806.

33. Harrop E, Mann M, Semedo L, et al. What elements of a systems’ approach to bereavement are most effective in times of mass bereavement? A narrative systematic review with lessons for COVID-19. Palliative medicine 2020; 34: 1165–1181.

34. Sheila A. Donahue MA, Nancy H. Covell PD, M. Jameson Foster MS, et al. Demographic Characteristics of Individuals Who Received Project Liberty Crisis Counseling Services. Psychiatric Services 2006; 57: 1261–1267. DOI: 10.1176/ps.2006.57.9.1261.

35. Koffman J, Donaldson N, Hotopf M, et al. Does ethnicity matter? Bereavement outcomes in two ethnic groups living in the United Kingdom. Palliat Support Care 2005; 3: 183-190. 2006/04/06. DOI: 10.1017/s1478951505050303.

36. Spruyt O. Community-based palliative care for Bangladeshi patients in east London. Accounts of bereaved carers. Palliative Medicine 1999; 13: 119-129. Research Support, Non-U.S. Gov’t.

37. Ackroyd R. Audit of referrals to a hospital palliative care team: role of the bilingual health-care worker. Int J Palliat Nurs 2003; 9: 352-357. 2003/09/12. DOI: 10.12968/ijpn.2003.9.8.11519.

38. Science and Technology Select Committee. Coronavirus: lessons learned to date. London: UK Government., 2021.

39. Public Health England. Beyond the data: Understanding the impact of COVID-19 on BAME groups 2020. UK Government.

40. Hussain JA, Koffman J and Bajwah S. Invited Editorial: Racism and palliative care. Palliative Medicine 2021; 35: 810-813. DOI 10.1177/02692163211012887.

