## Supplementary material for "“Sadly I think we are sort of still quite white, middle-class really” – Inequities in access to bereavement support: Findings from a mixed methods study": Suppl file 1 Survey

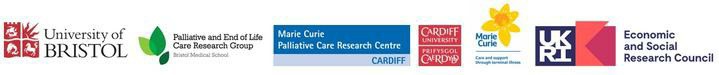

Bereavement During the COVID-19 Pandemic: National Survey of Bereavement Support Services

### Page 1: About this survey

Thank you for considering taking part in this survey. We appreciate that this is a busy time for all those working in bereavement support and are really grateful for your help.

This survey is part of a national study funded by the Economic and Social Research Council. We are working closely with the National Bereavement Alliance, and our findings will feed directly into policy and practice. We have already collected valuable data from around 700 people bereaved since 16 March 2020, and we want to represent the views and experiences of those providing bereavement services too. Thank you for your support.

##### WHO SHOULD COMPLETE THIS SURVEY?

This survey is for managers/coordinators/bereavement care leads of voluntary/community sector (VCS)* bereavement services in the UK. One person per organisation or branch (e.g. of Cruse) should complete it, **but please ask others in the organisation if you don’t have the information requested.** The survey is about:

The services you provide and how these have been affected by the pandemic How the pandemic has affected the demand and need for your services

How the pandemic has affected staff

The challenges you face as an organisation

To download or print a PDF of the survey to see the questions before you start, please click

###### here.

The information you provide will directly inform policy and practice. Interim findings from the study are publicly available at [www.covidbereavement.com.](http://www.covidbereavement.com/)

*By voluntary and community sector we mean non-profit, non-governmental organisations (e.g. charities, social enterprises, community groups). This does not include services provided solely by the NHS or commercial organisations, but does include palliative care or other bereavement services part-funded by the NHS. Please do not complete if your service is based within the NHS and receives no charitable funding. Eligible bereavement services could be stand-alone or embedded within a larger organisation.

##### ABOUT THIS SURVEY

**Background:** Bereavement is associated with increased risks to mental and physical health, which are likely to have increased during the COVID-19 pandemic when bereaved people have had less access to support from family and friends. Appropriate support following bereavement is essential, but bereavement services have faced many challenges during the pandemic and have had to adapt to difficult circumstances.

**Purpose:** This survey aims to understand the bereavement support available during the COVID-19 pandemic, how bereavement services and providers have been affected, and how services have responded to the challenges of the pandemic.

**Deadline:** This survey closes on 30th April 2021

##### DATA PROTECTION

Before you decide whether to consent to participate, please read the information below.

**Voluntary participation:** Participation is voluntary, you may choose not to answer survey questions unless these are required. If you decide to withdraw at any stage, you do not have to give a reason why.

**Confidentiality:** The information you provide will only be seen by the project staff who are undertaking this study. Any information we publish that might identify you, or others, will be anonymised.

**Data protection and usage:** All data collected in this survey will be held securely at the University of Bristol in accordance with the General Data Protection Regulations (GDPR; EU 2016/679). For more information on data protection, please follow the link: General Data Protection Regulation (GDPR; EU 2016/679). Cookies, personal data stored by your web browser, are not used in this survey.

By participating in this survey, you are agreeing that your survey responses and anonymised extracts of text may be included in future reports, professional journals and future research.

The results will be disseminated via newsletters, blogs, conferences, peer-reviewed journal articles, press releases, social media and the study website [[www.covidbereavement.com](http://www.covidbereavement.com/)]. The anonymised data will be archived and might be used in secondary analysis or for educational purposes in the future. Your personal data (identifying information) will be kept securely for 10 years after the research is completed in line with University of Bristol policies in

addition to GDPR.

**Risks and benefits:** When using the internet, there can be a risk of compromising privacy, confidentiality and/or anonymity. We are using a secure platform to conduct the survey to minimise these risks. By completing this survey, you are helping understand the bereavement support currently available and how bereavement services have been affected by and responded to the challenges of the COVID-19 pandemic.

**Contact information:** If you have any concerns or questions relating to this survey, please contact Dr. Eileen Sutton, Senior Research Associate at or 07977021723

Additionally, if you have a concern about the conduct of the study, please contact Dr. Lucy Selman at Palliative and End of Life Care Research Group, University of Bristol, Canynge Hall, 39 Whatley Road, Bristol BS8 2PS. If you have a complaint about the study, please contact the University’s independent Research Governance team by emailing Complaints or concerns are taken seriously and will be addressed immediately.

##### INSTRUCTIONS FOR COMPLETION

One representative from each bereavement support organisation (or branch of an organisation) is eligible to take part. This will usually be a paid member of staff (e.g. manager, service lead) but could be a volunteer.

Please ask others in your organisation for any information requested which you don’t have yourself.

The survey should take 20-30 minutes to complete.

Please add any comments in the blank boxes that you feel are relevant or important. Feel free to use bullet points.

The survey is designed to capture information on many different types of services and some questions may appear less relevant to you. We would be grateful if you could please try to answer all questions that apply.

You can save your responses and return to complete your survey later. To do so, please click on the 'finish later' link at the bottom of the page. You will then need to either bookmark your unique URL link in your browser or ask for it to be emailed to you.

You can navigate the survey and edit your answers up to the point you click the 'finish survey' button. To go back in the survey (e.g. to revise a response), please click the ‘previous’ button at the bottom of the page.

CONSENT: By agreeing to participate in this survey, you indicate that you have read and understood the information above and that you are aged 18 or over.

**Consent:** I consent to participate in this survey and I agree that I have read and understood the information provided above.  *Required*

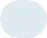

*1.*

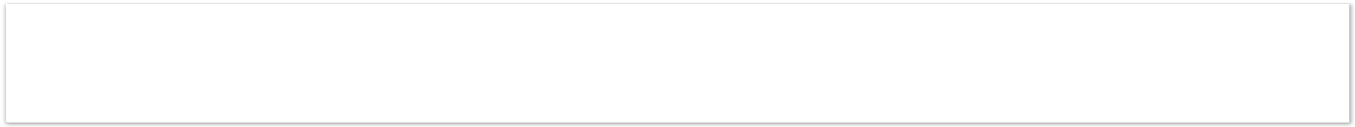

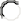

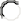

Yes No

### Page 2: DEMOGRAPHICS

#### About you

Job title:  *Required*

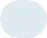

*2.*

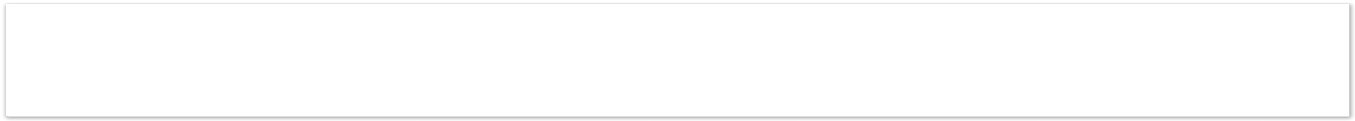

Name of your bereavement service:  *Required*

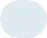

*3.*

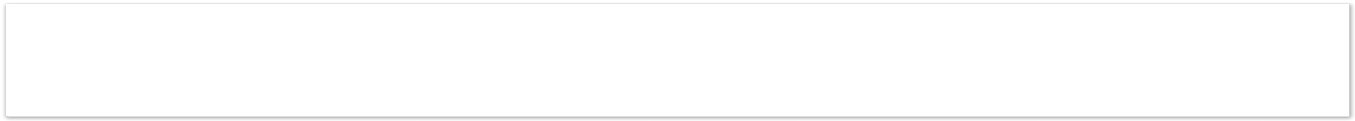

If your bereavement service is a local branch of a national organisation/charity (e.g.

*3.a.*

Cruse, SOBs, MIND, Age UK, Place2Be), please state the name of that organisation:

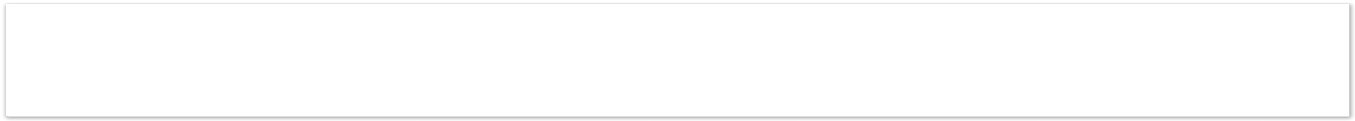

#### PART A: ABOUT YOUR SERVICE

What kind of catchment area do you serve?  *Required*

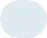

*4.*

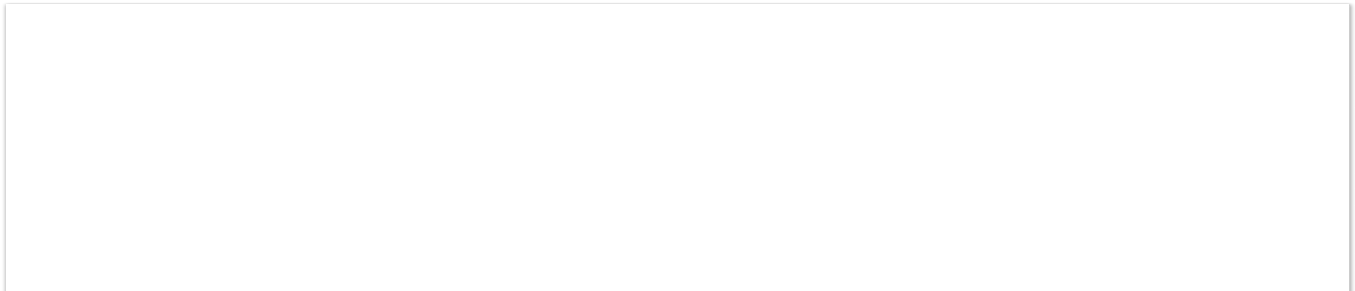

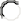

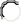

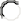

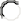

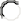

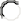

UK-wide

Nation-wide: England only Nation-wide: Wales only Nation-wide: Scotland only

Nation-wide: Northern Ireland only

Specific Counties/Local Authorities/Clinical Commissioning Groups (CCGs) - please

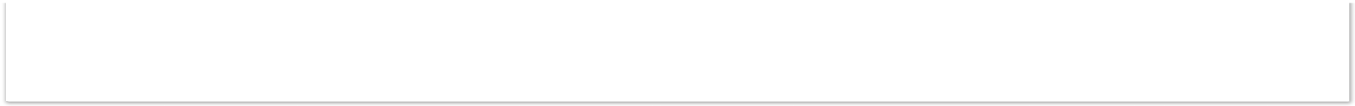

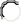

specify below:

Other specific catchment area - please specify below:

If you selected "Specific Counties/Local authorities/CCG Area", please specify:

*4.a.*

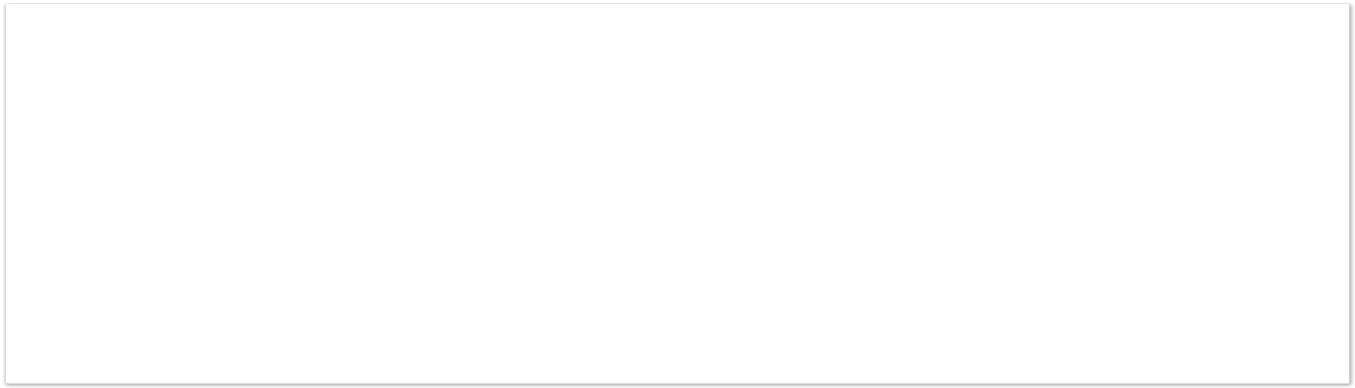

If you selected "Other Specific Catchment Area", please specify:

*4.b.*

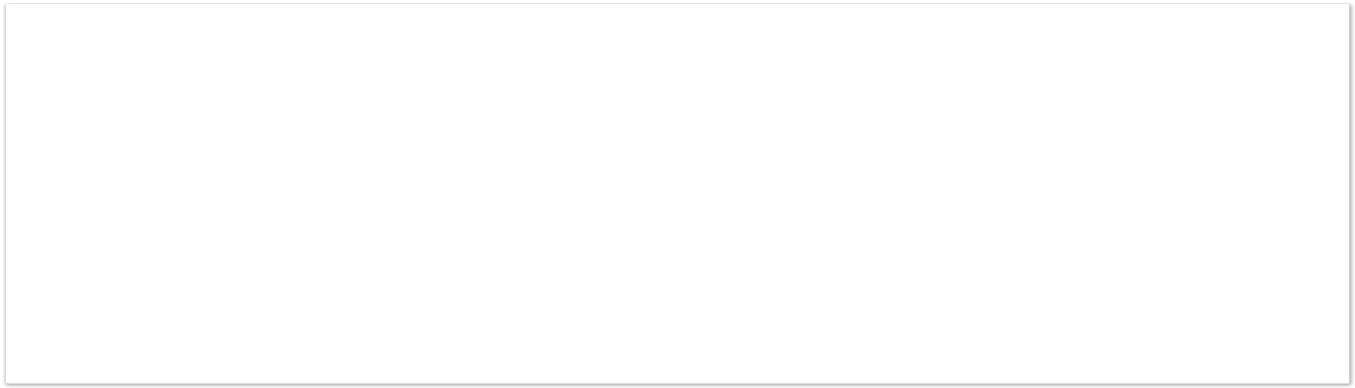

Which of the following best describes your organisation:  *Required*

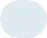

*5.*

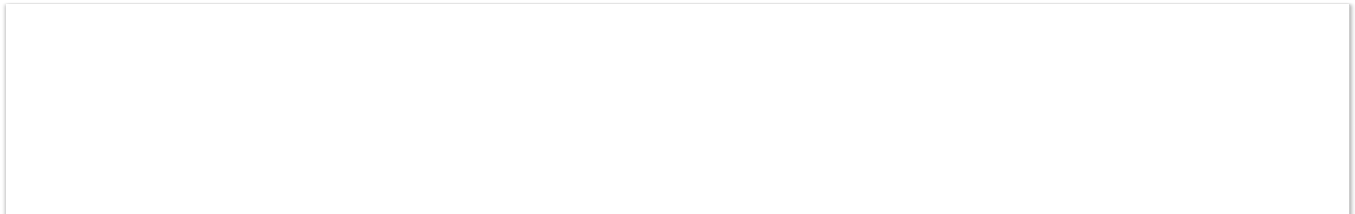

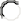

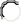

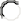

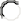

National bereavement charity / Non-governmental Organisation (NGO) Local bereavement charity / NGO

Branch of national bereavement charity / NGO (e.g. SOBS area, Cruse branch) Branch of other national charity / NGO (e.g. MIND, Age UK, Place2Be, Marie Curie)

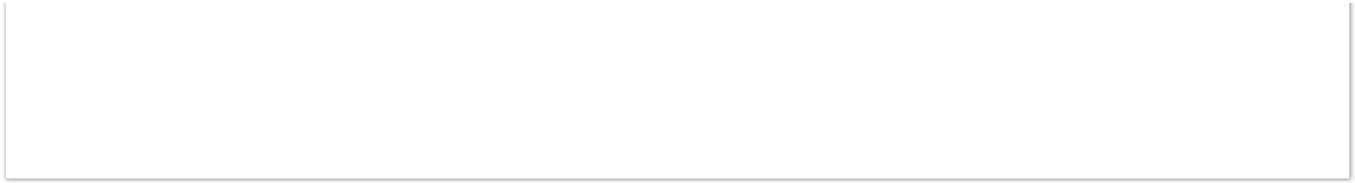

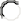

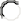

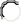

Hospice/palliative care team bereavement service part-funded by NHS (Please do not complete if your service receives no charitable funding)

Local other charity / NGO (e.g. generic counselling service) Other - please specify below:

If you selected "Other", please specify:

*5.a.*

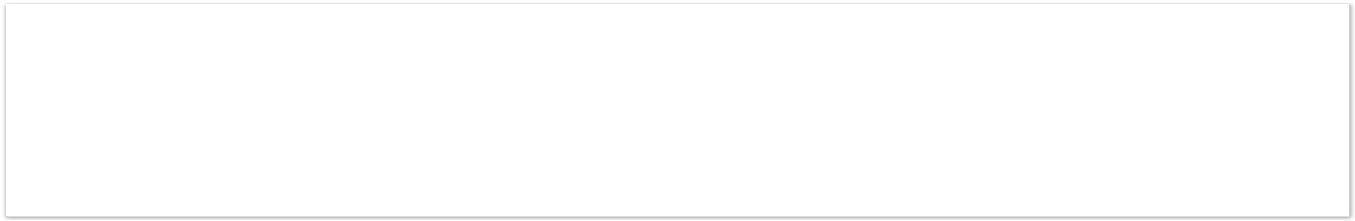

What age groups are the primary focus of your work? Tick all that apply:  *Required*

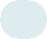

*6.*

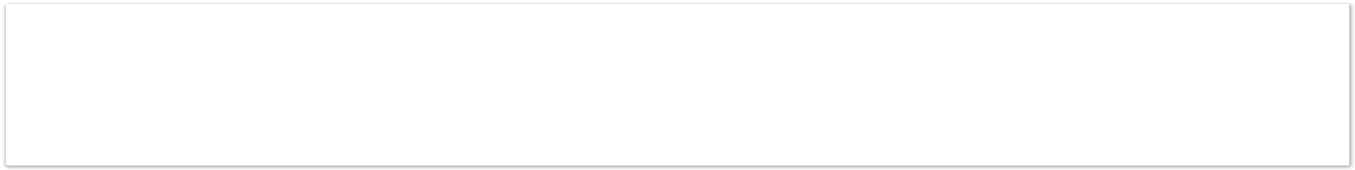

Bereaved children and young people (and those caring for them) Bereaved adults

Other - please specify below:

If you selected "Other" please specify:

*6.a.*

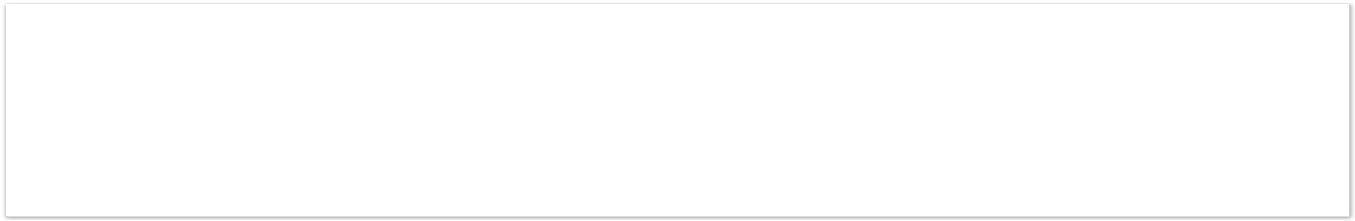

Is the support that you offer restricted to people already known to your organisation? (e.g. family members of deceased patients cared for by your service)  *Required*

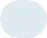

*7.*

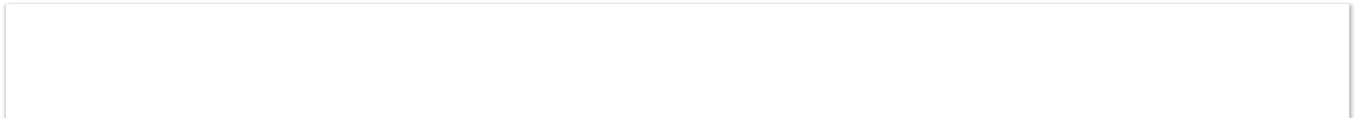

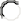

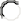

Yes No

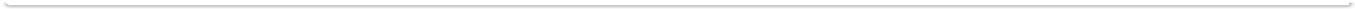

Is the support that you offer restricted to any other group of people not listed above (e.g. members of particular religious/cultural group, particular society or institution etc):  *Required*

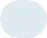

*8.*

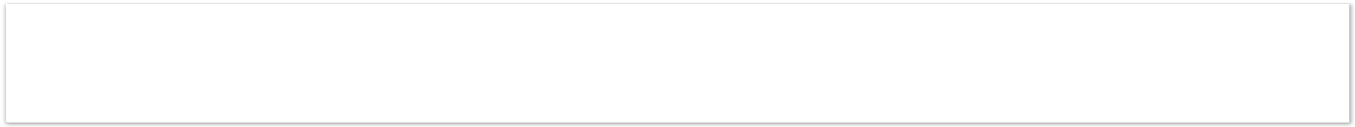

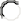

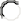

Yes No

If you selected "Yes", please specify:

*8.a.*

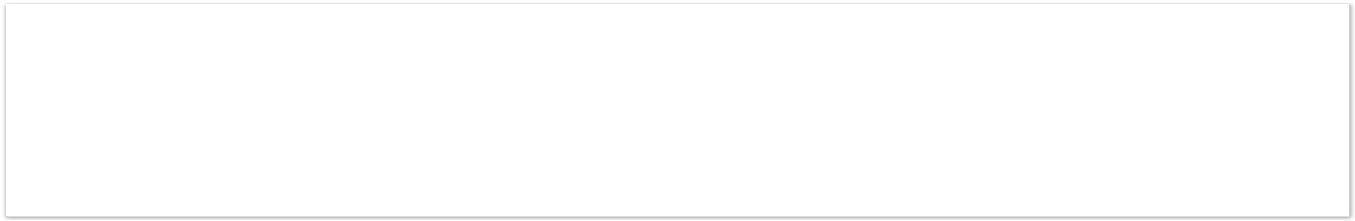

Do you specialise in providing support according to the age of the person who died? 

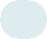

*9.*

*Required*

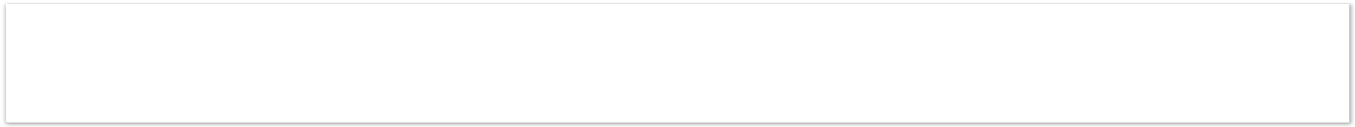

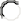

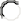

Yes No

If you selected "Yes", please tick all that apply

*9.a.*

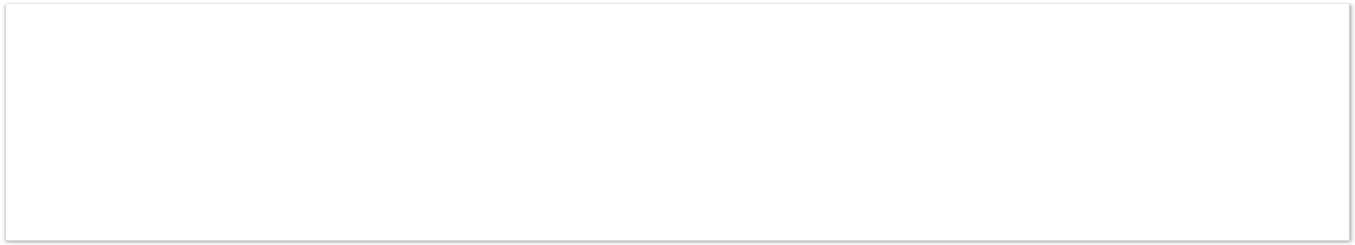

Loss during pregnancy (including miscarriage, ectopic/ molar pregnancy, termination of pregnancy for any reason)

Stillbirth and/or neonatal death

The death of a child or young person Adult death

Following what causes of death does your organisation support bereaved people? 

*10.*

*Required*

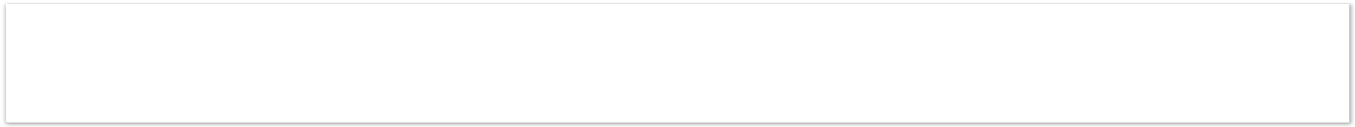

Following all causes of death

Following specific causes of death (e.g. life-limiting illness, suicide, cancer)

If you selected "Specific causes of death", please specify:

*10.a.*

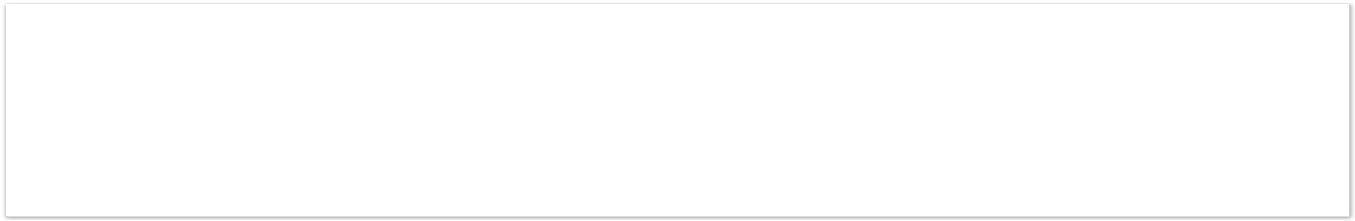

#### Services provided pre-Coronavirus

Please indicate which services you provided **before COVID-19** and how you provided them. Please tick all that apply:  *Required*

*11.*

|  |  | Online | Via |
| --- | --- | --- | --- |
|  | Face to face | (website,  web chat, Skype, | telephone  (helpline, mobile |
|  |  | Zoom) | app) |
| Information on grief and sign-posting to other services |  |  |  |
| Group meeting of peers (people with similar experiences but no one is trained) |  |  |  |
| Group meeting facilitated by someone with training |  |  |  |
| One-to-one support (e.g. individual or family counselling by someone with training) |  |  |  |
| Specialist intervention involving mental health services, psychological support services or specialist counselling/psychotherapy |  |  |  |

Did your organisation provide any other bereavement services **before COVID-19?**

*12.*

Please tick all that apply: *Optional*

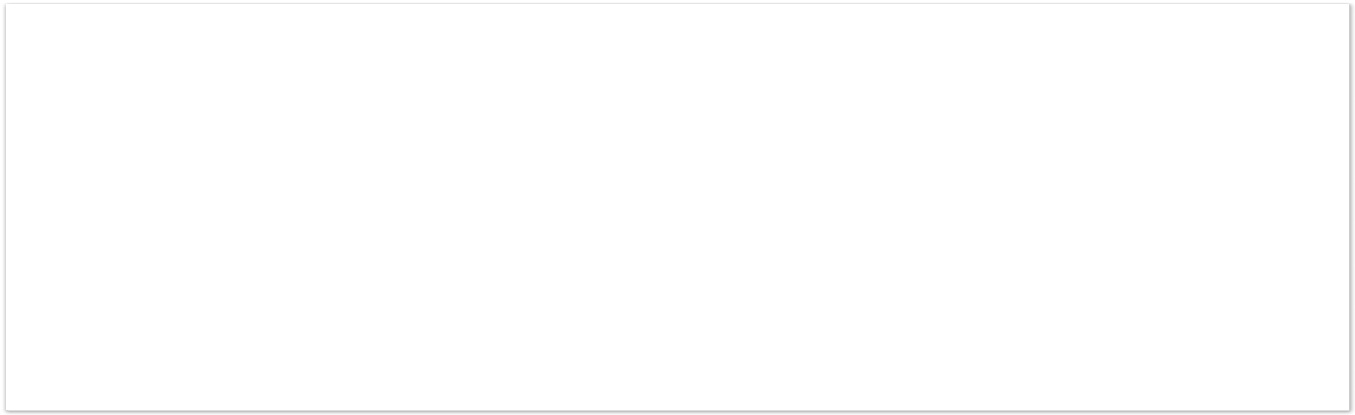

Pre-death support/bereavement preparation (e.g. memory-making activities) Immediate post-death support (e.g. bereavement suites, memory making activities) -

please specify below: Condolence letters Home visit Remembrance services Drop-in support

Online community

Other - please specify below:

If you indicated that you provide immediate post-death support, please briefly describe

*12.a.*

this:

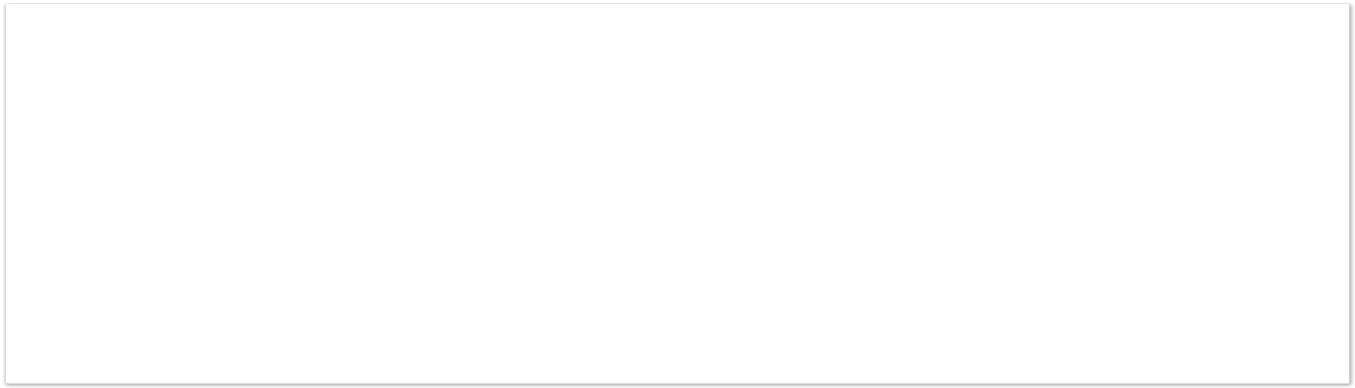

If you selected "Other", please specify:

*12.b.*

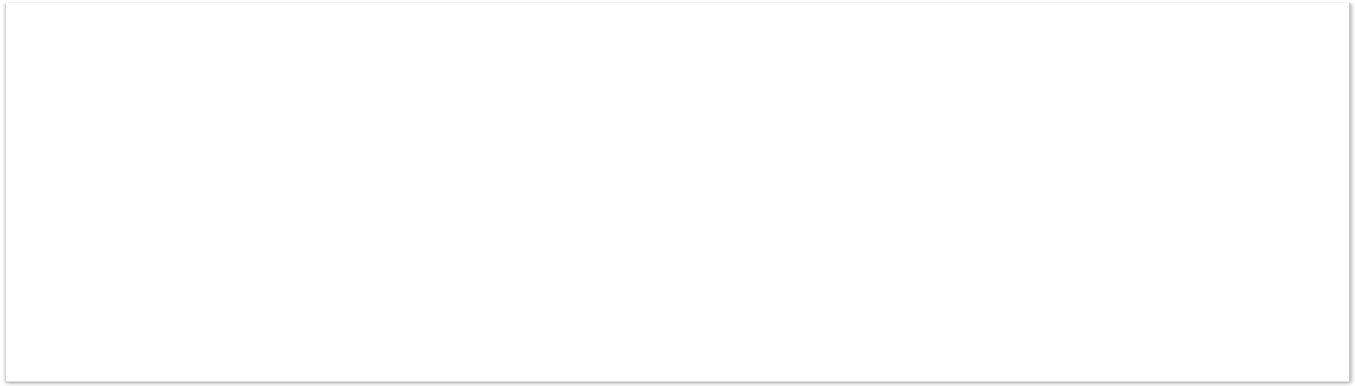

###### The following questions ask about the clients supported by your service before COVID-

**19. Please ask another member of staff if you don’t have the information yourself.**

Before COVID-19, please estimate how many clients your service would **usually** support each month? (If your service only collects annual data, please divide your annual figure by 12)

*13.*

 *Required*

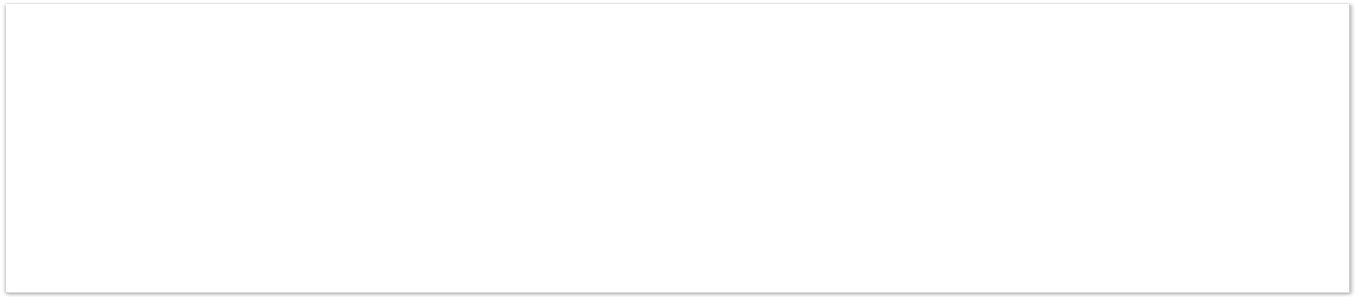

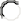

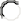

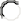

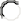

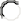

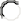

Up to 20

21 - 40

41 - 60

61 - 80

81 or over

I don't know

**In the year before COVID-19**, what proportion of your clients were from Black or minority ethnic communities? Please select your closest estimate.  *Required*

*14.*

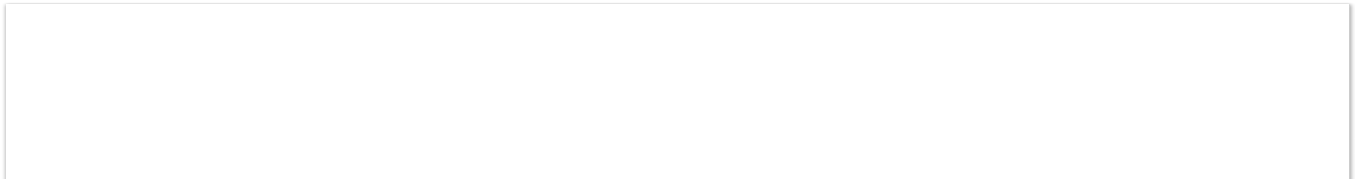

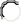

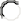

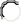

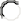

Less than 5%

5 - 10%

11 - 15%

16 - 20%

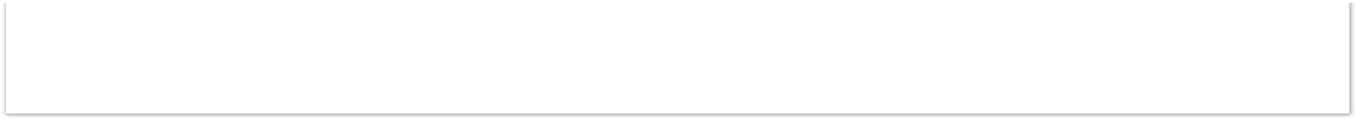

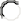

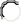

More than 20%

We don't collect this data

Before COVID-19, do you think there were specific community groups with unmet bereavement support needs that you were not reaching, or who experienced barriers to accessing your service?  *Required*

*15.*

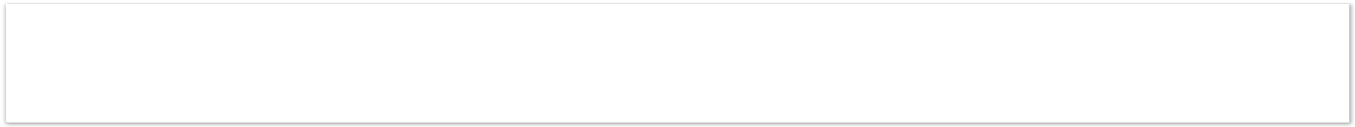

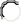

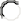

Yes No

If you selected "Yes" which groups? Please tick all that apply

*15.a.*

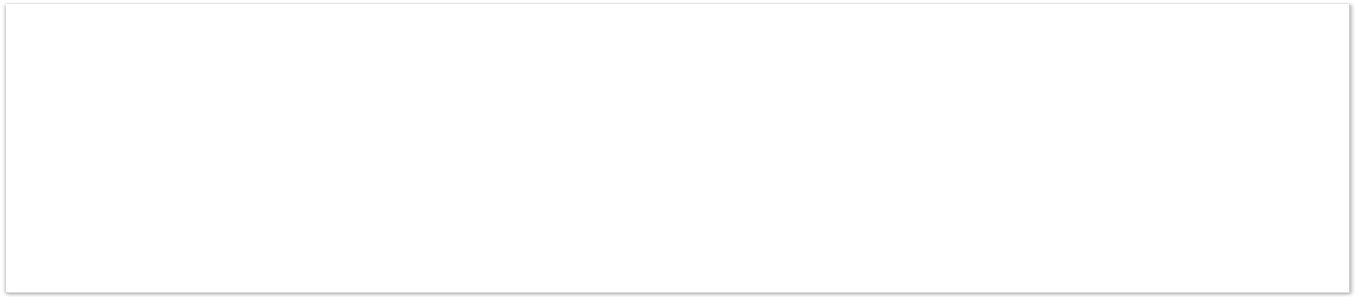

People from lower socioeconomic groups/poorer areas Men

Younger people

People from ethnic minority groups People from sexual minority groups Other (please state below)

If you selected "Other", please specify:

*15.a.i.*

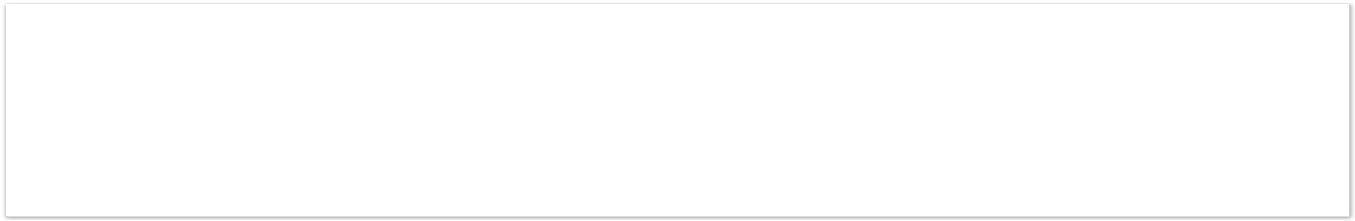

### Page 3: PART B: IMPACT OF COVID-19

#### Impact on Service Delivery

Which of these services are you **currently** providing? Please tick all that apply: 

*16.*

*Required*

|  |  | Online | Via |
| --- | --- | --- | --- |
|  | Face to face | (website, web chat,  Skype, | telephone (helpline,  mobile |
|  |  | Zoom) | app) |
| Information on grief and sign-posting to other services |  |  |  |
| Group meeting of peers (people with similar experiences but no one is trained) |  |  |  |
| Group meeting facilitated by someone with training |  |  |  |
| One-to-one support (e.g. individual or family counselling by someone with training) |  |  |  |
| Specialist intervention involving mental health services, psychological support services or specialist counselling/psychotherapy |  |  |  |

Is your organisation providing any other bereavement services **now?** Please tick all that apply: *Optional*

*17.*

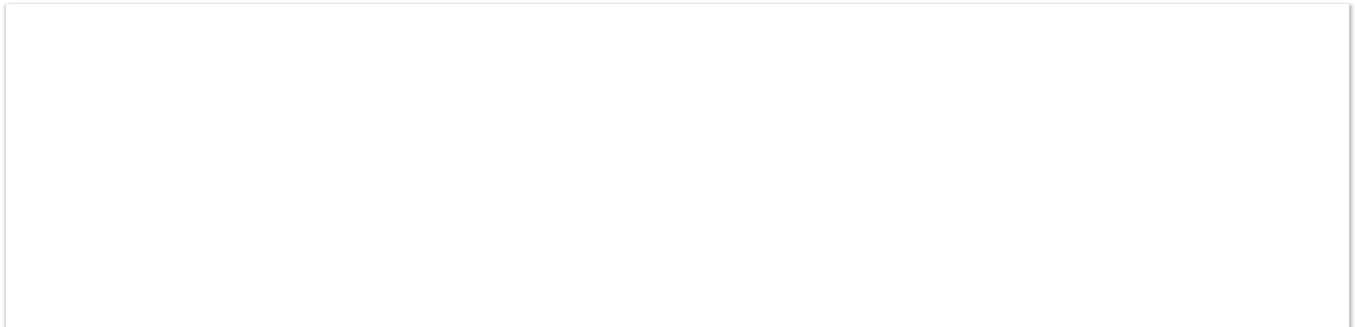

Pre-death support/bereavement preparation (e.g. memory-making activities) Immediate post-death support (e.g. bereavement suites, memory making activities) Condolence letters

Home visit Remembrance services Drop-in support

Online community

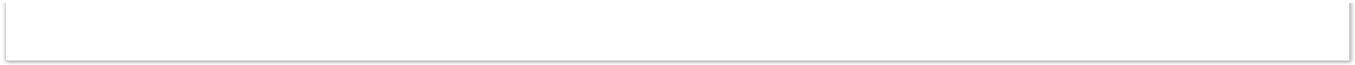

Other - please specify below:

If you selected "Other", please specify:

*17.a.*

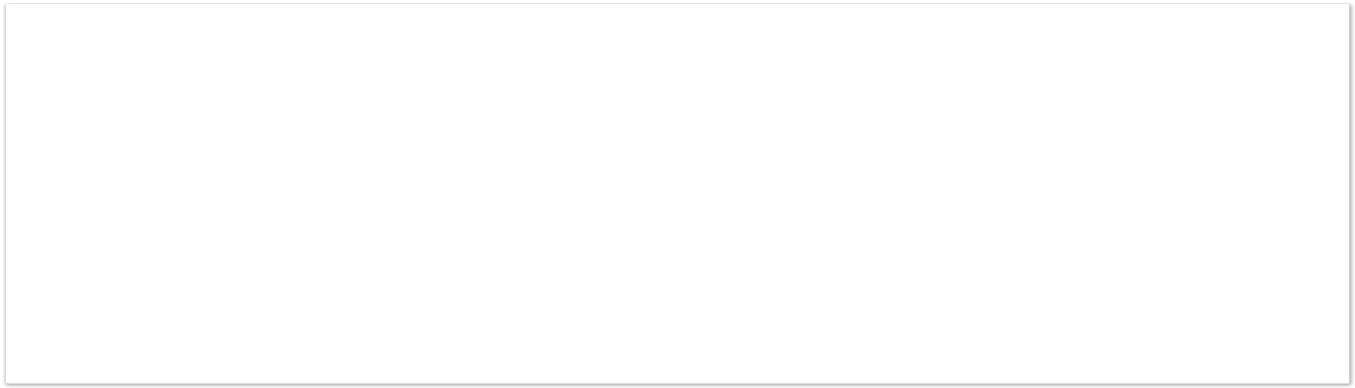

How has COVID-19 affected the support you provide? Please tick all that apply: 

*18.*

*Required*

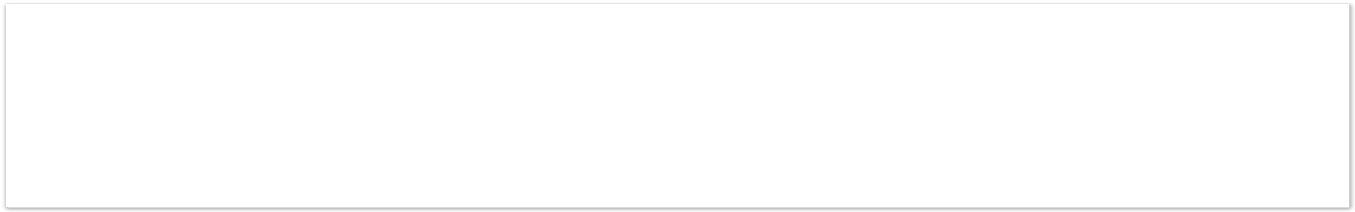

We stopped delivering services completely

We changed existing services (please describe below) We introduced new services (please specify below)

We made other changes to our services or support (please describe below)

If you indicated that you changed existing services, please describe these changes:

*18.a.*

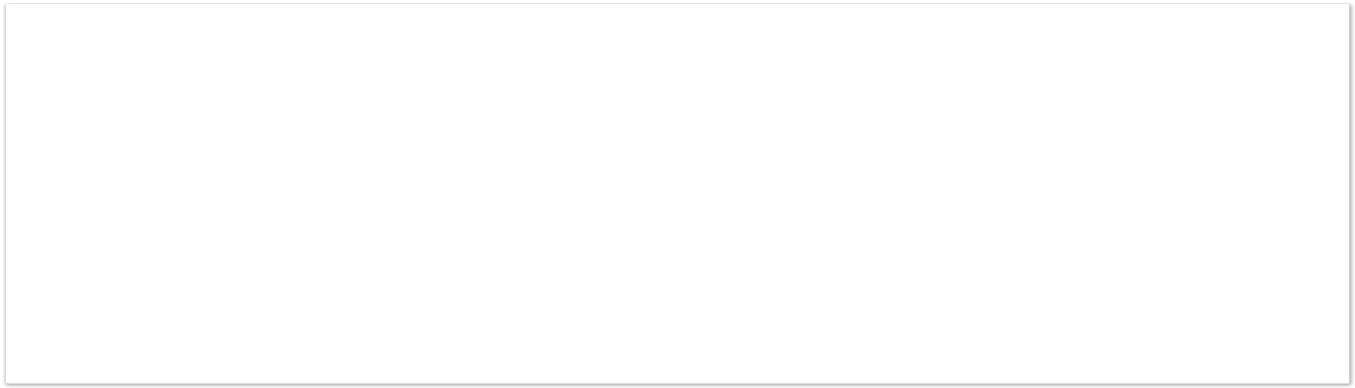

If you indicated that you introduced new services, please specify:

*18.b.*

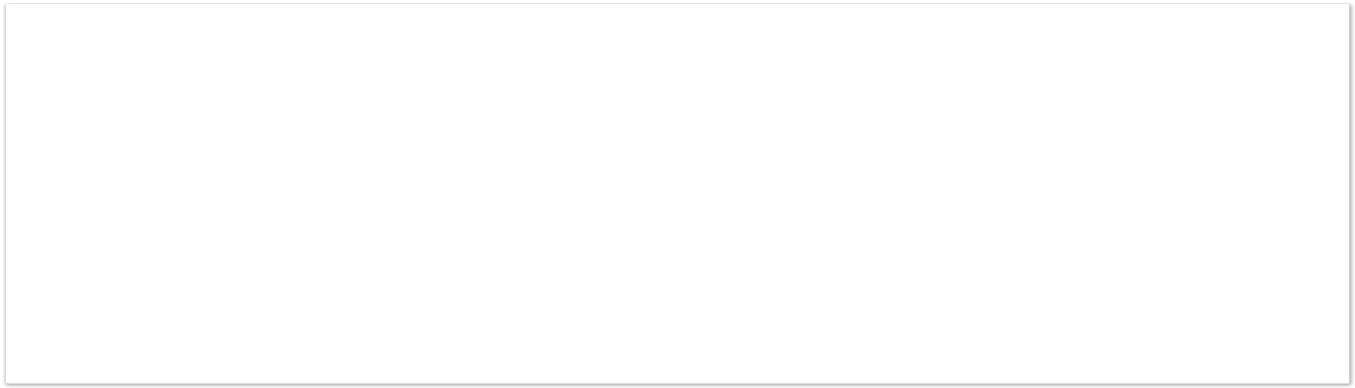

If you indicated that you made other changes to your services or support, please describe these changes:

*18.c.*

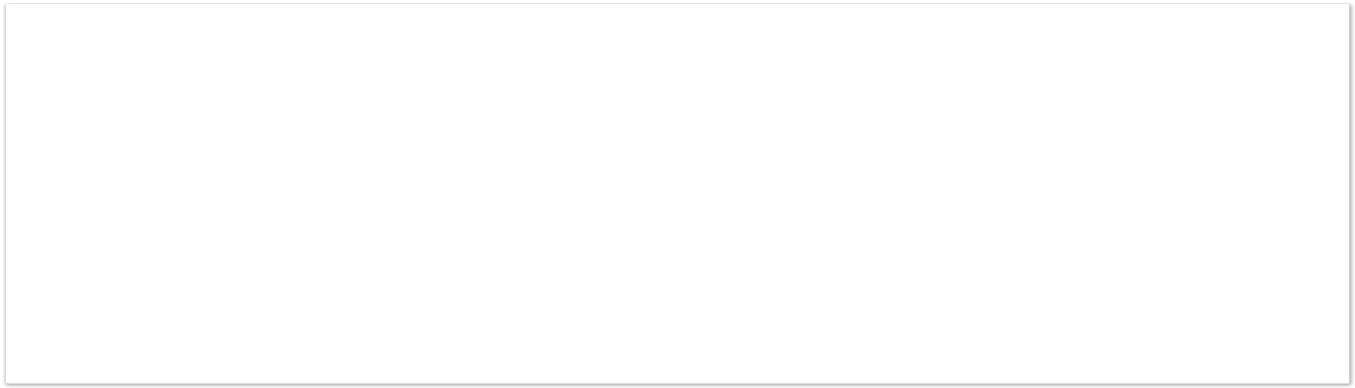

Do you think COVID-19 has led to any positive changes or opportunities for your service?  *Required*

*19.*

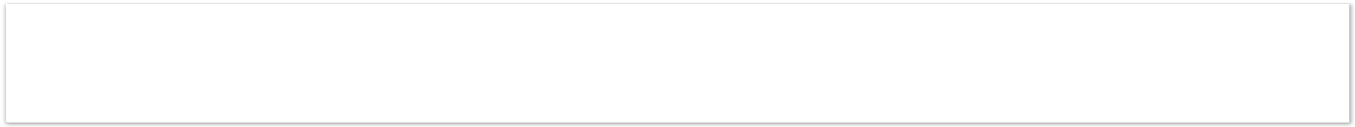

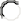

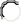

Yes No

If you selected "Yes", please tell us about any positive changes or opportunities:

*19.a.*

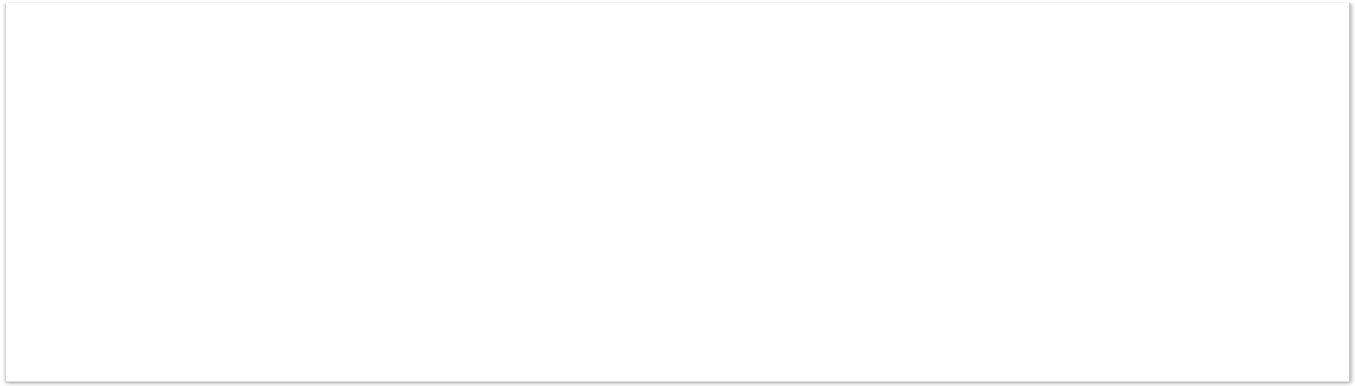

Is there anything else you would like to say about how COVID-19 has impacted on the services you have provided since March 2020? For example, are the changes you made during the first lockdown ongoing? Have any changes in services impacted on your ability to meet clients’ needs?

*20.*

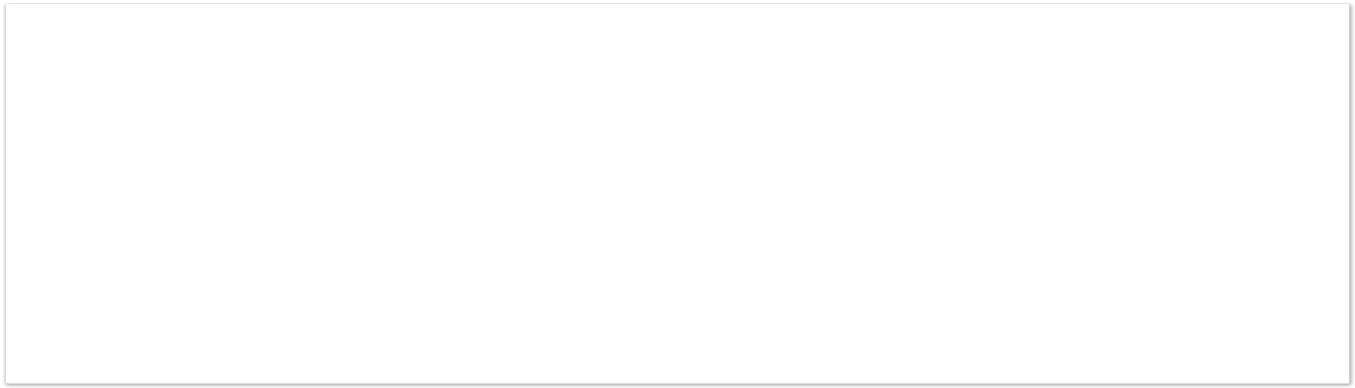

We are interested in identifying examples of good and innovative practice in how bereavement services have offered support to clients during the pandemic, which other services might be able to learn from. If you have any examples, please describe below:

*21.*

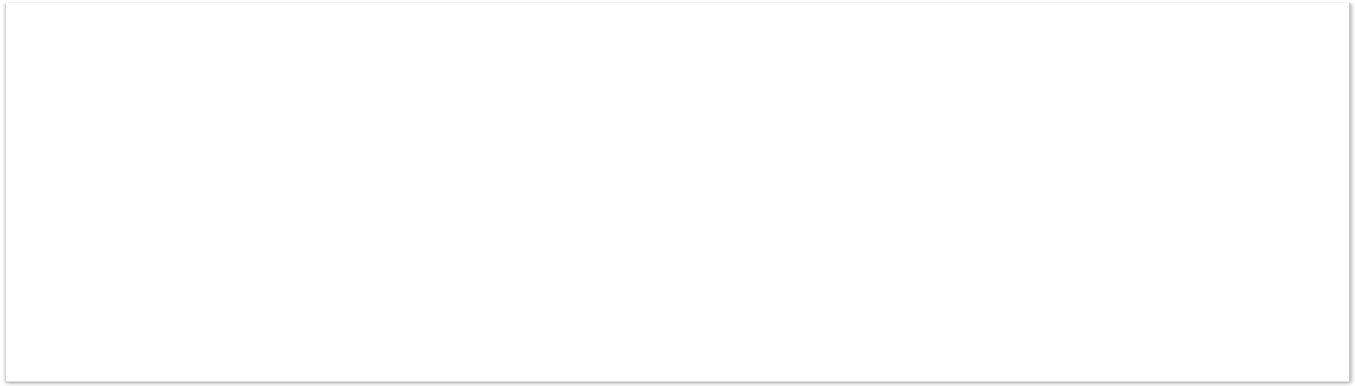

We are also interested in changes in practice which have been especially challenging to implement or not worked so well. If you have any examples, please describe below:

*22.*

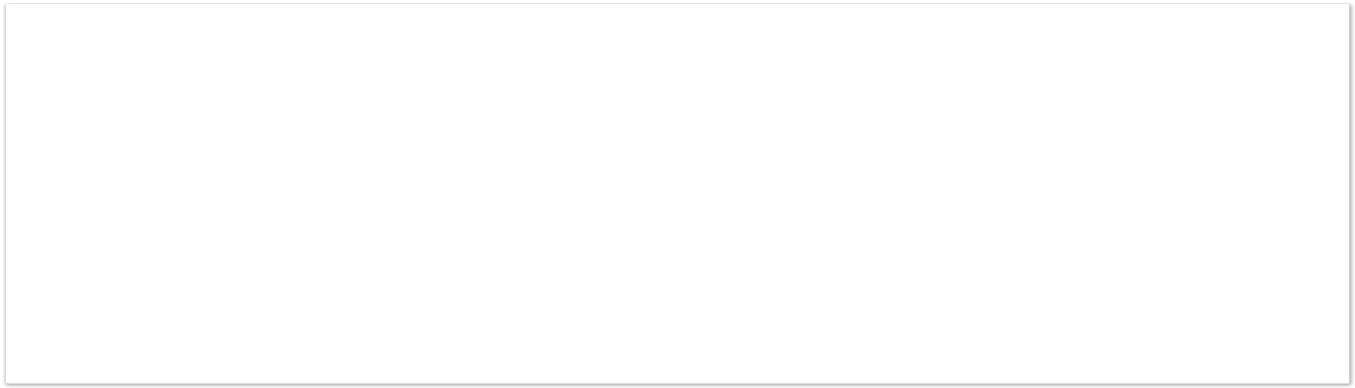

#### Impact on Service Uptake and Client Needs

Since March 2020, what has happened to your referrals compared to what you would expect in a usual year?  *Required*

*23.*

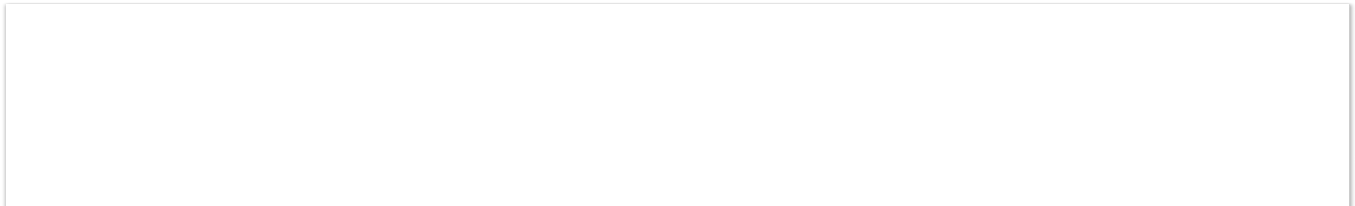

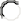

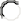

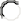

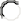

Much lower than usual (more than 25% lower) A bit lower than usual (up to 25% lower) About the same as usual

A bit higher than usual (up to 25% higher)

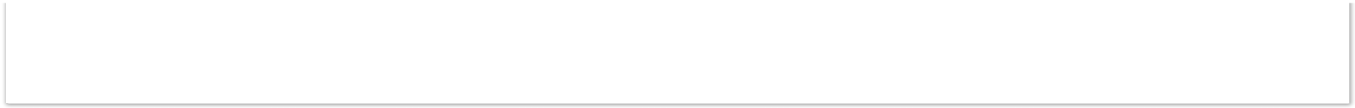

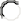

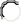

Much higher than usual (more than 25% higher) I don't know

If you can, please describe these patterns of referral in more detail (for example “the number of people wanting to join our support groups dropped by over 50% early in the first lockdown, but it is now back to usual”, or “the number of people ringing our helpline

*24.*

increased by 20% early in the first lockdown, and then by another 20% over July and August, and has stayed at this rate”).

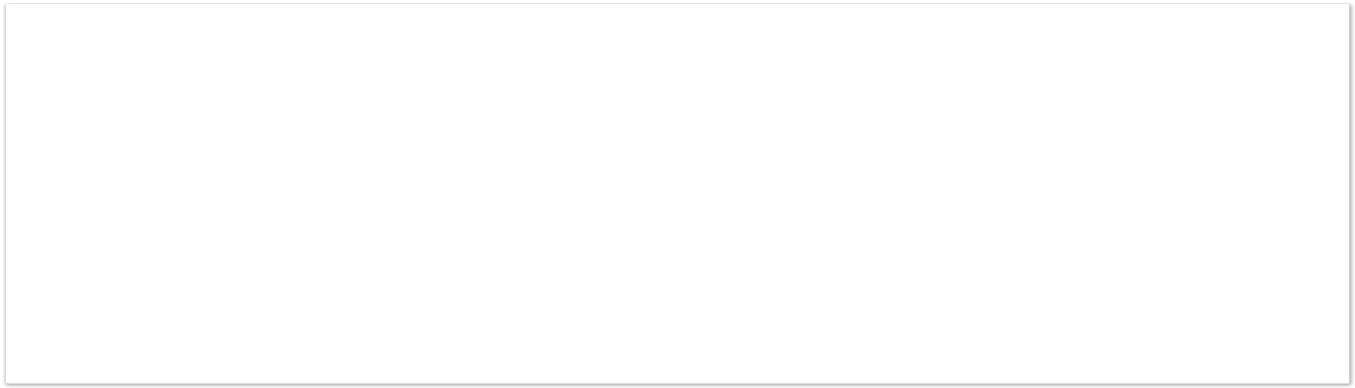

Compared with before COVID-19, has the proportion of clients you support from Black and minority ethnic communities changed since March 2020?  *Required*

*25.*

More of our current clients are from Black or other minority ethnic groups Approximately the same proportion

Fewer of our current clients are from Black or other minority ethnic groups I don't know

We don't collect this data

Has the way you advertise or accept referrals to your service, or the way that people hear about your service changed since March 2020? Please describe below.  *Required*

*26.*

Is there anything else you would like to tell us about the number or type of people accessing your services? Please use the box below:

*27.*

How has COVID-19 affected the bereavement support needs of your clients? For example, are you seeing more complex needs or higher levels of risk? If relevant to your service, please also comment on clients’ pre-bereavement support needs. Please answer in the box below:  *Required*

*28.*

#### Impact of COVID-19 on the Workforce

How did COVID-19 affect paid staff involved in providing bereavement support at your service? Please tick all that apply:  *Required*

*29.*

Staff were furloughed

Staff were made redundant Higher levels of staff illness

Fewer staff able to work for other reasons e.g. caring responsibilities Staff providing bereavement support from home

Increased pressure on staff due to volume of clients Increased pressure on staff due to nature of client needs Increased emotional impact on staff

Increased need for supervision/support Difficulties recruiting staff

Other - please specify below:

Not applicable - no effect on paid staff

If you selected "Other", please specify:

*29.a.*

How did COVID-19 affect volunteers involved in providing bereavement support at your service? Please tick all that apply:  *Required*

*30.*

The service has stopped volunteers from working Higher levels of volunteer sickness

Fewer volunteers able to work for other reasons e.g. caring responsibilities Volunteers providing bereavement support from home

Increased pressure on volunteers due to volume of clients Increased pressure on volunteers due to nature of clients Increased emotional impact on volunteers

Increased need for volunteer supervision/support Difficulties recruiting volunteers

Other - please specify below:

Not applicable - no effect on our volunteers Not applicable - we don't work with volunteers

If you selected "Other", please specify:

*30.a.*

If you would like to say more about the effects on paid staff or volunteers, please use the box below:

*31.*

Did COVID-19 affect staff training at your service?  *Required*

*32.*

Yes No

In what ways did COVID-19 affect staff training? Please tick all that apply:

*32.a.*

Staff training courses were cancelled or postponed Staff training was delivered online

Staff were trained on COVID-19 hygiene measures

Staff were trained on providing adapted bereavement support/counselling during COVID-19 (e.g. online counselling)

Staff were trained on meeting the needs of people bereaved during the pandemic, including COVID-19 deaths

Other - please specify below:

If you selected "Other", please specify:

*32.a.i.*

#### Gaps and Challenges in Service Provision

Do you currently face financial challenges in service delivery?  *Required*

*33.*

Yes No

What kind of financial challenges do you currently face in service delivery? Please tick all that apply:

*33.a.*

Fundraising events have been cancelled or postponed Reduced income due to organisational cuts in spending Less access to corporate funding

Less access to funds from Trusts and foundations

Less access to funding from individual dontations

Restrictions related to COVID-19 grants (e.g. competition for grants, narrow criteria, restrictions to type or timeframe of expenditure) – please specify below:

Other financial challenges - please specify below:

If you indicated that you have faced restrictions related to COVID-19 grants, please

*33.a.i.*

specify:

If you selected "Other financial challenges", please specify:

*33.a.ii.*

Does your service currently face any non-financial challenges?  *Required*

*34.*

Yes No

What kind of non-financial challenges do you currently face? Please tick all that apply:

*34.a.*

Access to pandemic-specific training to support our clients

Access to other training - please describe any specific gaps below: Access to specialist support to refer on to e.g. mental health services

Access to appropriate facilities e.g. COVID-secure space for face-to-face support Lack of volunteers able to work

Unable to meet demand for service Unable to meet client needs

Other - please specify below:

If you indicated that you experienced challenges in access to other training, please describe any specific gaps:

*34.a.i.*

If you selected "Other", please specify:

*34.a.ii.*

Is there currently a waiting list to access any aspects of your service?  *Required*

*35.*

Yes - less than three weeks

Yes - between 3 weeks and 2 months Yes - between 2 months and 4 months Yes - between 4 months and 6 months Yes - more than 6 months

No

Please tell us more about the challenges your organisation faces, either now or in the future, in the box below:

*36.*

Please tell us more about anything that has helped you with your work through the pandemic:

*37.*

Have there been any local/regional initiatives to coordinate bereavement services in your area?  *Required*

*38.*

Yes No

If you selected "Yes" please tell us more about these initiatives:

*38.a.*

### Page 4: What next?

Would you be happy to be contacted by a researcher to take part in a telephone interview to discuss in more detail how your service has been affected by, or responded to, the pandemic? Interviews will take place at a mutually convenient time and take approximately 30 minutes.  *Required*

*39.*

Yes - please complete your contact details below: No

Name:

*39.a.*

Work email address:

*39.b.*

Work contact telephone number:

*39.c.*

Please enter a valid phone number.

The study website [www.covidbereavement.com](http://www.covidbereavement.com/) reports regular updates on the progress of this study. If you would also like to receive a summary of the results of this study, please provide your email address here. Your address will not be shared beyond the study team. If you have any further questions, please contact Dr. Eileen Sutton, Senior Research Associate at or 07977021723

*40.*

### Page 5: Thank you

**Thank you very much for completing this survey.** Your answers will help ensure the bereaved and bereavement services in the UK are adequately supported.
