## Supplementary material for "“Sadly I think we are sort of still quite white, middle-class really” – Inequities in access to bereavement support: Findings from a mixed methods study": Suppl file 2 Topic guide

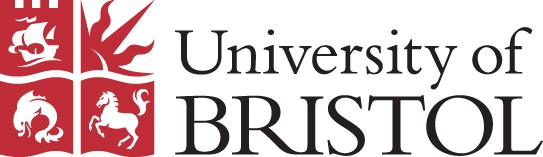

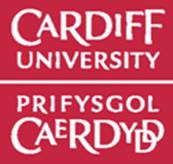

#### The impact of the COVID-19 pandemic on bereavement support services

**Topic Guide for interviews with bereavement service providers: Version 1 18/11/2020**

**NOTE:** This is an outline of topics explored – revised in line with emerging findings and examples of innovation from online survey. This topic guide was adapted for online bereavement support providers.

### 1) INTRODUCTION

Introduction, explanation of study aims and audio recording permission Audio consent taking

2) **PARTICIPANT BACKGROUND** (Brief discussion to contextualise responses) Organisation/location/geographical coverage

Type of support provided by organisation

Role within organisation/length/depth of experience/previous role

#### 3) IMPACT OF COVID-19 Changing demand and level of support

Changes to requests for support (probe: across different times/stages of pandemic)

- Whether or not demand has increased
- Any increased demand from BAME communities or from lower socio-economic groups/more marginalised communities
- Timing of support request (e.g. earlier, later)

Level of support needed:

- More complex grief experiences?
- Tier 1 support (probe: relative numbers, demand for information provision)
- Tier 2 support (probe: relative numbers, individual and group support)
- Tier 3 support (probe: relative numbers, specialist mental health and psychological support, waiting lists for referral)

#### Changes to therapeutic encounter

Impact of providing support remotely

- Loss of non-verbal communication (probe: bereaved person, provider)
- Changes in support relationship and impact
- Changes in interaction with those seeking support and impact (probe: characteristics of bereaved person – age, gender, ethnicity, socio-economic status etc)
- IT/practical issues
- How they have coped with these changes

#### Impact on staff providing support

Coping with increased/changing demand

- Feeling unprepared
- Managing increased demand?
- Managing increased number of people with complex/complicated grief responses?
- Waiting lists
- Ability to do their job as they would like
- Staff illness/shielding
- Personal support/supervision for staff/volunteers
- How they have coped with these changes

### 4) INNOVATION/CHANGE

**Innovations and/or changes in delivery** (NOTE: questions developed further using examples provided in survey)

Services withdrawn/on hold New/extended/innovative services

- Adapting existing services
- New services provided
- Responding quickly to increased demand
- COVID-specific services
- Reaching different groups

How changes came about

- Why changes needed
- Who instigated changes
- Who was involved in designing/developing changes
- Who was involved in providing the services
- How the changes were implemented (probe: funding, technology, training, skills)
- COVID-specific challenges
- Ways of meeting the challenges faced
- What worked well/what didn’t work – what were the keys to success? Or the reasons for difficulties?
- Any additional support that would have been helpful
- What would they do differently (probe: lessons for other providers)
- What were the positives (probe: reaching different groups, targeting services)

Evaluating services

- Have they carried out evaluation of new/changed services
- If yes - how did they evaluate, any results (probe; published, given feedback)
- If not - any difficulties (probe: time, support needed)
- Short or long-term changes anticipated (probe; changing Covid landscape)

### 5) RESOURCES/MANAGING CHANGE

Impact of increased demand on staffing Loss of volunteers (shielding etc)

Negative impact on funding streams (probe: fundraising, restrictions related to COVID-specific funding)

#### Training/support needs of staff/volunteers

Types of training

- Dealing with complex/complicated grief responses
- Delivering support remotely
- Individual/group delivery
- IT/practical support
- Personal support/supervision

### ANYTHING ELSE

How do they view provision moving forward

Opportunity for participant to add anything else important to them not covered

Suggestions for additional key participants within their organisation for further interviews – i.e. to give a different perspective (e.g. staff vs volunteer, fundraising vs service delivery) or to help complete our understanding of the service and its response to the pandemic

**THANKS**
