## Supplementary material for "“Sadly I think we are sort of still quite white, middle-class really” – Inequities in access to bereavement support: Findings from a mixed methods study": Suppl file 3 Tables

**Table I. Results of logistic regression for there being specific community groups with unmet**

**bereavement support needs not being reached/experiencing barriers to**

**accessing your service (yes or no)**

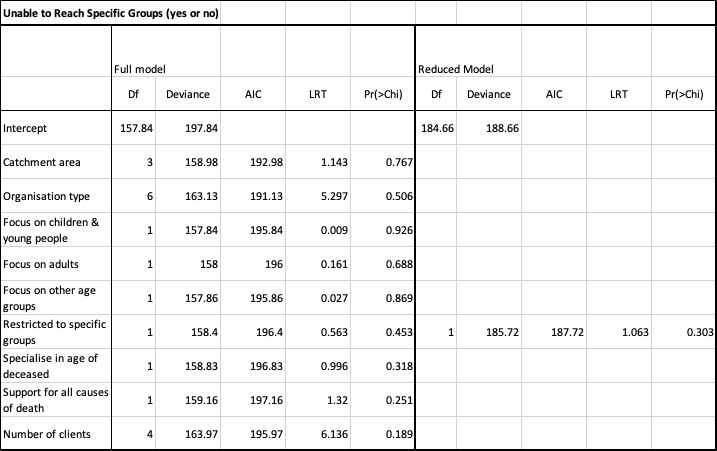

**Table II. Results of logistic regression for proportion of clients from minoritised ethnic groups (<5% or ≥5%)**

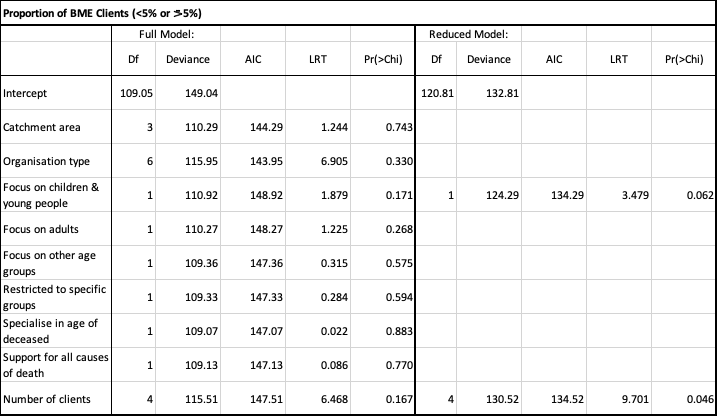

**Table III. Results of logistic regression for collection of ethnicity data (yes/no)**

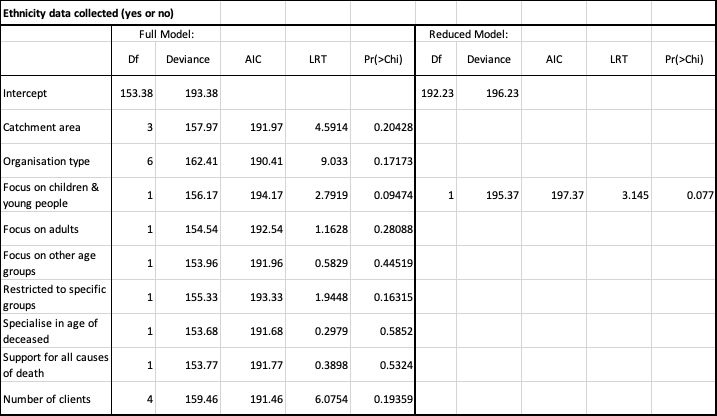
